# A locally deployable agentic framework for clinical data deidentification

**DOI:** 10.64898/2026.05.28.26353952

**Authors:** Anja Hirsch, Foo Wei Ten, Kyle S. Krüger, Robin C. Geyer, Tobias Röschl, Matthias I Gröschel, Paul Rostin, Roland Eils, Martin Spott, Fabian Prasser, Alexander Meyer, Julian Madrid

## Abstract

Multimodal clinical data contain identifiers across diverse formats. We developed the Multimodal Anonymizer, a locally deployable framework combining multimodal language models, specialist networks, deterministic transformations and iterative verification. We compared 16 orchestrator configurations using automated evaluation on 250 constructed MIMIC-IV-derived patient bundles with synthetic identifiers. A selected local configuration underwent manual evaluation on these same bundles supplemented with head CT scans, face images, handwritten notes, audio recordings and synthetic German clinical notes from unrelated sources. Separate evaluations used the German GRASCCO corpus and 250 Charité partograms. On this expanded benchmark, patient-level deidentification sensitivity was 98.80% (95% CI, 97.20–100), per-identifier sensitivity 99.82% (95% CI, 99.76–99.88), patient-level critical clinical preservation 99.60% (95% CI, 98.80–100) and the clinical preservation score 99.61% (95% CI, 99.51–99.71). All partograms met the study-defined deidentification criterion. These findings support feasibility, while constructed bundles, synthetic identifiers, limited natural-data validation and absent component ablations constrain generalizability.

## Introduction

Artificial Intelligence (AI) is transforming many fields and is increasingly reshaping health care. Multimodal models that combine text, images, waveforms, laboratory values, and longitudinal clinical events offer the prospect of substantial quality and efficiency improvements^1,2^.

To realize that potential, multimodal models must be trained on large, diverse, high-quality datasets^3^. Yet an estimated 97% of hospital data goes unused, and privacy constraints around patient-identifying information are an important barrier to reuse^4,5^.

Current approaches address only parts of this problem. Named-entity recognition can remove identifiers from text^6^ and deep-learning methods can detect sensitive content in individual modalities^7^, but these methods are typically modality-specific^5^ and often have limited contextual understanding^8^—a central requirement when identifiers are embedded across different types of real-world clinical data. Context-enhanced deidentification systems have been developed^9^, and local large language models have shown promise for deidentifying medical text^10^. However, integrating contextual deidentification across heterogeneous clinical data remains challenging.

The next generation of clinical AI will require deidentification systems that are multimodal, scalable, locally deployable, privacy-preserving, and yet simple enough for people without coding expertise to use. Different deidentification aspects can benefit from different computational strategies: multimodal large language models (MLLMs) for contextual reasoning, specialized deep-learning models for modality-specific detection, and rule-based methods for deterministic transformations^11^. Agentic workflows combine the advantages of all three strategies while mitigating their respective limitations^11,12^.

We developed and retrospectively evaluated a modular system for multimodal deidentification that can run locally within institutional environments and can be accessed through a web-based user interface. We benchmarked the system against existing approaches and evaluated performance through deidentification sensitivity and critical clinical preservation. Our primary hypothesis was that such a system could deidentify medical data without over-redacting clinically critical information. Secondary hypotheses were that system performance would improve with orchestrator size, reasoning capability, full-precision deployment, and dense architectures; that the integrated workflow would outperform existing approaches; and that performance would remain stable across repeated runs.

## Results

### Primary Results

Following automated evaluation on DeID-Core, our MIMIC-IV-derived benchmark with synthetically injected identifiers, Qwen3-VL-235B-A22B-Thinking was selected for detailed manual evaluation on DeID-Extended, which supplements DeID-Core with additional modalities. Two independent domain experts reviewed all deidentified outputs, assessing both completeness of identifier removal and preservation of clinically relevant content.

The Multimodal Anonymizer achieved per-patient and per-PII (personally identifiable information) deidentification sensitivities of 98.80% (95% CI, 97.20–100) and 99.82% (95% CI, 99.76–99.88), respectively (Figure 1a). For all ten modality groups, lower 95% confidence bounds for sensitivity were at least 98.30% (Figure 1b); sensitivity on the separate GRASCCO corpus was 96.77% (95% CI, 94.70–98.61).

**Figure 1:**
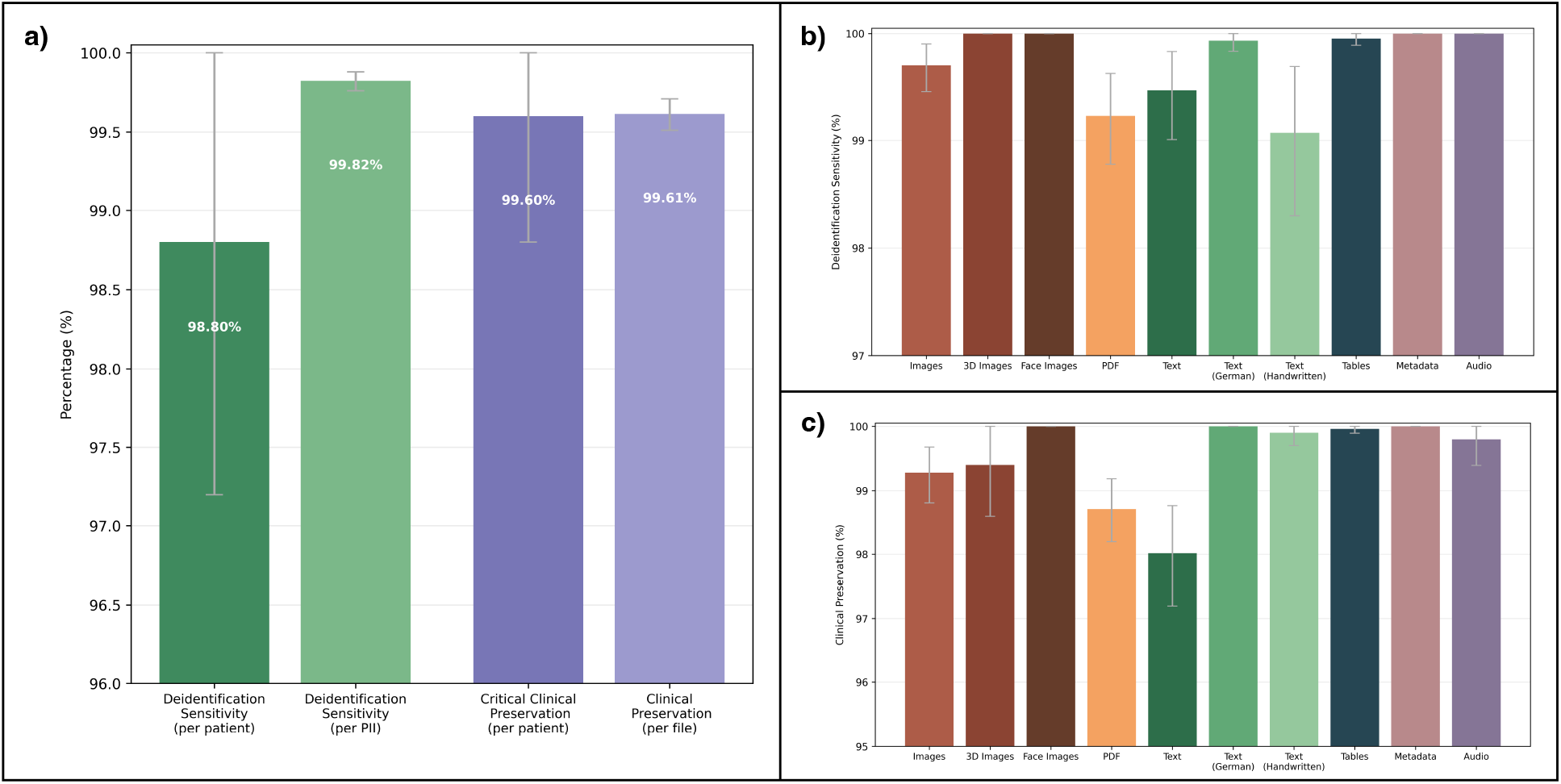
Manual evaluation results of deidentification sensitivity, critical clinical preservation, and clinical preservation score on DeID-Extended using Qwen3-VL-235B-A22B-Thinking. Deidentification sensitivity and (critical) clinical preservation are reported at multiple aggregation levels. Deidentification sensitivity is reported per-patient (proportion of patients without any critical omission) and per-PII entity (proportion of all identifier entities across the ten modality groups that were adequately redacted). Patient-level deidentification sensitivity reflects whether any single residual identifier or a combination of residual identifiers across a patient’s files could enable reidentification; a patient is counted as adequately deidentified only if no such identifier or combination remains (see Supplementary Table S2). Critical clinical preservation denotes the proportion of patients for whom all highly clinically relevant non-PII content pertaining to the current case, such as diagnoses, medications, and treatment plans, remained unaltered after deidentification. The clinical preservation score is derived from file-level redaction annotations and calculated as described in Statistical Analysis. All panels show 95% confidence intervals. **Panel a)** shows overall deidentification sensitivity, critical clinical preservation and the clinical preservation score. **Panel b)** shows per-PII deidentification sensitivity stratified by modality, and **panel c)** shows the per-file clinical preservation score stratified by modality. CI denotes confidence interval; and PII, personally identifiable information.

Critical clinical preservation was 99.60% (95% CI, 98.80–100) at the patient level (Figure 1a). A single instance of clinically critical information loss was identified across DeID-Extended (one active pharmaceutical ingredient was redacted from a discharge note). The clinical preservation score based on file-level redaction annotations was 99.61% (95% CI, 99.51–99.71), with lower 95% confidence bounds of at least 97.19% across all modalities (Figure 1c). Full results are available in Supplementary Table S1.

For site-specific validation, we performed an identical analysis on the Charité partogram dataset, comprising records from 250 patients. The system achieved a deidentification sensitivity of 100% at both per-patient and per-PII aggregation levels, with no recorded study-defined failures (Table 1). Critical clinical preservation was likewise 100% per-patient, with no clinically critical information redacted. The clinical preservation score based on file-level redaction annotations was 99.97% (95% CI, 99.91–100).

**Table 1.**
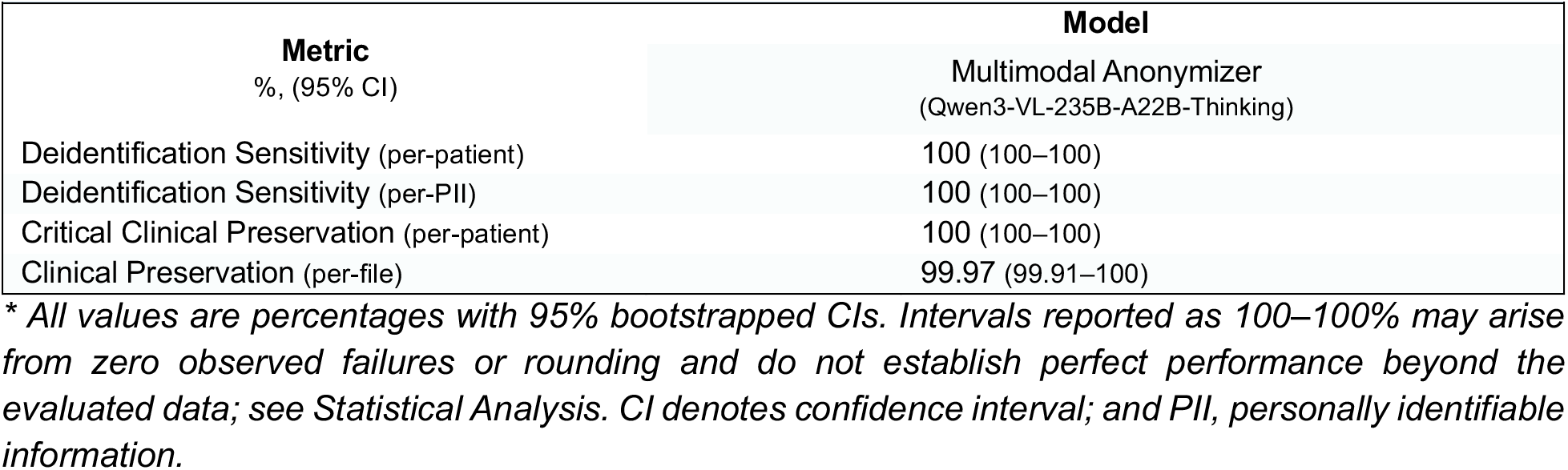
Deidentification Sensitivity, Critical Clinical Preservation and Clinical Preservation Score on the Charité partogram dataset.*

### Secondary Results

Complete automated evaluation metrics on DeID-Core and results of pairwise model comparisons, with between-model differences (Δ) expressed in percentage points (pp), are provided in Supplementary Table S3 and Figs. S1–S2.

#### Performance across models

Table 2 summarizes automated deidentification performance on DeID-Core for five representative high-capacity models (see Supplementary Figs. S3–S6 for visualizations). Overall deidentification sensitivity scores were highest for GPT-5.2, followed closely by Qwen3.5-397B-A17B-thinking and Qwen3-VL-235B-A22B-Thinking. GPT-5.2 showed slightly higher deidentification sensitivity than Qwen3.5-397B-A17B-thinking (Δ = 0.06 pp; 95% CI, 0.03 to 0.10) and Qwen3-VL-235B-A22B-Thinking (Δ = 0.07 pp; 95% CI, 0.01 to 0.16), although the magnitude of these differences was small. GPT-5.2 achieved higher results than Kimi-K2.5-thinking (Δ = 1.01 pp; 95% CI, 0.65 to 1.41) or GLM-4.6V (Δ = 1.08 pp; 95% CI, 0.43 to 2.04).

**Table 2.**
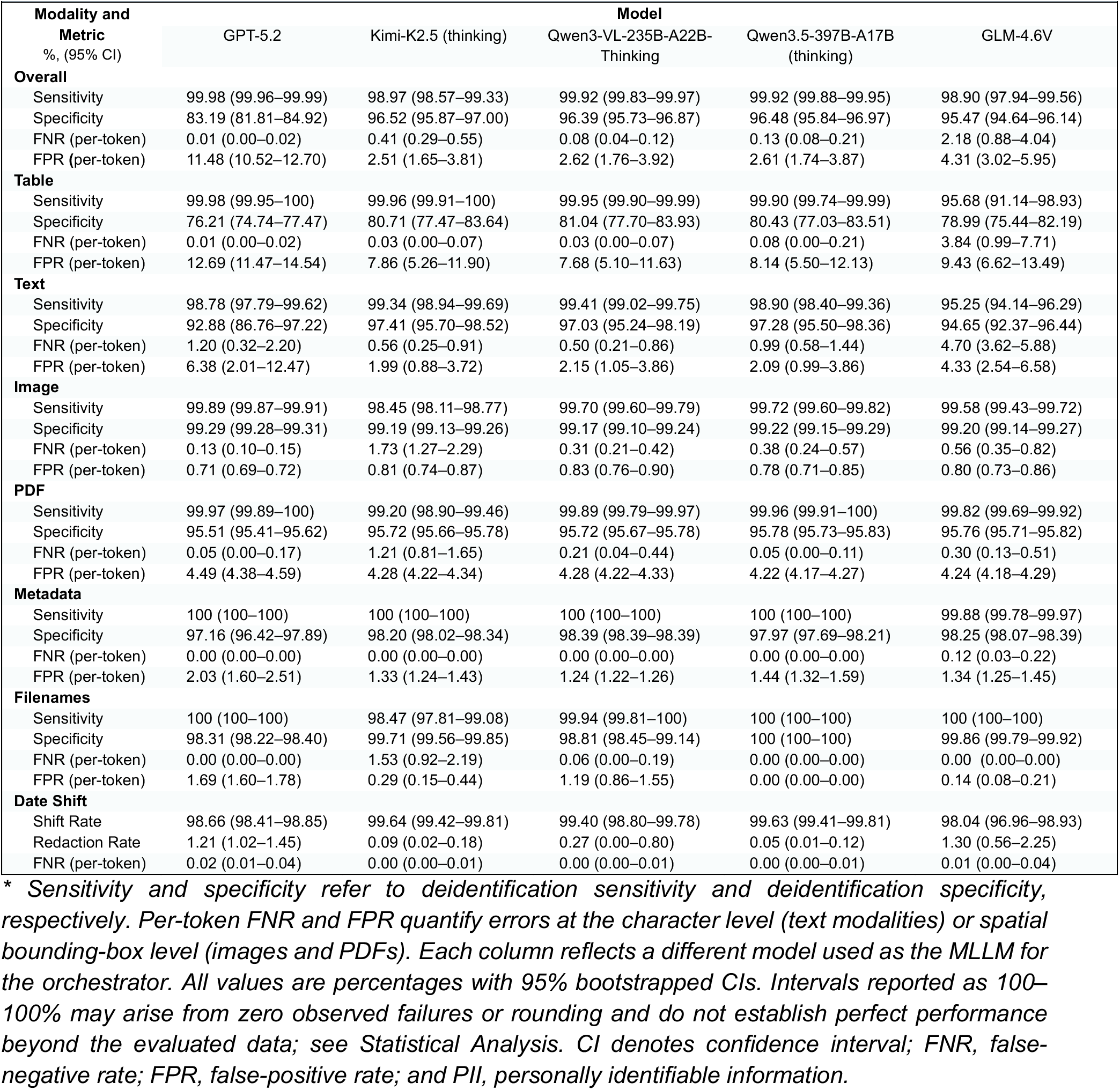
Automated Evaluation of Deidentification Performance on DeID-Core.*

| **Modality and Metric**<br>%, (95% CI) | **GPT-5.2** | **Kimi-K2.5 (thinking)** | **Qwen3-VL-235B-A22B-Thinking** | **Qwen3.5-397B-A17B (thinking)** | **GLM-4.6V** |
| --- | --- | --- | --- | --- | --- |
| **Overall** |  |  |  |  |  |
| Sensitivity | 99.98 (99.96–99.99) | 98.97 (98.57–99.33) | 99.92 (99.83–99.97) | 99.92 (99.88–99.95) | 98.90 (97.94–99.56) |
| Specificity | 83.19 (81.81–84.92) | 96.52 (95.87–97.00) | 96.39 (95.73–96.87) | 96.48 (95.84–96.97) | 95.47 (94.64–96.14) |
| FNR (per-token) | 0.01 (0.00–0.02) | 0.41 (0.29–0.55) | 0.08 (0.04–0.12) | 0.13 (0.08–0.21) | 2.18 (0.88–4.04) |
| FPR (per-token) | 11.48 (10.52–12.70) | 2.51 (1.65–3.81) | 2.62 (1.76–3.92) | 2.61 (1.74–3.87) | 4.31 (3.02–5.95) |
| **Table** |  |  |  |  |  |
| Sensitivity | 99.98 (99.95–100) | 99.96 (99.91–100) | 99.95 (99.90–99.99) | 99.90 (99.74–99.99) | 95.68 (91.14–98.93) |
| Specificity | 76.21 (74.74–77.47) | 80.71 (77.47–83.64) | 81.04 (77.70–83.93) | 80.43 (77.03–83.51) | 78.99 (75.44–82.19) |
| FNR (per-token) | 0.01 (0.00–0.02) | 0.03 (0.00–0.07) | 0.03 (0.00–0.07) | 0.08 (0.00–0.21) | 3.84 (0.99–7.71) |
| FPR (per-token) | 12.69 (11.47–14.54) | 7.86 (5.26–11.90) | 7.68 (5.10–11.63) | 8.14 (5.50–12.13) | 9.43 (6.62–13.49) |
| **Text** |  |  |  |  |  |
| Sensitivity | 98.78 (97.79–99.62) | 99.34 (98.94–99.69) | 99.41 (99.02–99.75) | 98.90 (98.40–99.36) | 95.25 (94.14–96.29) |
| Specificity | 92.88 (86.76–97.22) | 97.41 (95.70–98.52) | 97.03 (95.24–98.19) | 97.28 (95.50–98.36) | 94.65 (92.37–96.44) |
| FNR (per-token) | 1.20 (0.32–2.20) | 0.56 (0.25–0.91) | 0.50 (0.21–0.86) | 0.99 (0.58–1.44) | 4.70 (3.62–5.88) |
| FPR (per-token) | 6.38 (2.01–12.47) | 1.99 (0.88–3.72) | 2.15 (1.05–3.86) | 2.09 (0.99–3.86) | 4.33 (2.54–6.58) |
| **Image** |  |  |  |  |  |
| Sensitivity | 99.89 (99.87–99.91) | 98.45 (98.11–98.77) | 99.70 (99.60–99.79) | 99.72 (99.60–99.82) | 99.58 (99.43–99.72) |
| Specificity | 99.29 (99.28–99.31) | 99.19 (99.13–99.26) | 99.17 (99.10–99.24) | 99.22 (99.15–99.29) | 99.20 (99.14–99.27) |
| FNR (per-token) | 0.13 (0.10–0.15) | 1.73 (1.27–2.29) | 0.31 (0.21–0.42) | 0.38 (0.24–0.57) | 0.56 (0.35–0.82) |
| FPR (per-token) | 0.71 (0.69–0.72) | 0.81 (0.74–0.87) | 0.83 (0.76–0.90) | 0.78 (0.71–0.85) | 0.80 (0.73–0.86) |
| **PDF** |  |  |  |  |  |
| Sensitivity | 99.97 (99.89–100) | 99.20 (98.90–99.46) | 99.89 (99.79–99.97) | 99.96 (99.91–100) | 99.82 (99.69–99.92) |
| Specificity | 95.51 (95.41–95.62) | 95.72 (95.66–95.78) | 95.72 (95.67–95.78) | 95.78 (95.73–95.83) | 95.76 (95.71–95.82) |
| FNR (per-token) | 0.05 (0.00–0.17) | 1.21 (0.81–1.65) | 0.21 (0.04–0.44) | 0.05 (0.00–0.11) | 0.30 (0.13–0.51) |
| FPR (per-token) | 4.49 (4.38–4.59) | 4.28 (4.22–4.34) | 4.28 (4.22–4.33) | 4.22 (4.17–4.27) | 4.24 (4.18–4.29) |
| **Metadata** |  |  |  |  |  |
| Sensitivity | 100 (100–100) | 100 (100–100) | 100 (100–100) | 100 (100–100) | 99.88 (99.78–99.97) |
| Specificity | 97.16 (96.42–97.89) | 98.20 (98.02–98.34) | 98.39 (98.39–98.39) | 97.97 (97.69–98.21) | 98.25 (98.07–98.39) |
| FNR (per-token) | 0.00 (0.00–0.00) | 0.00 (0.00–0.00) | 0.00 (0.00–0.00) | 0.00 (0.00–0.00) | 0.12 (0.03–0.22) |
| FPR (per-token) | 2.03 (1.60–2.51) | 1.33 (1.24–1.43) | 1.24 (1.22–1.26) | 1.44 (1.32–1.59) | 1.34 (1.25–1.45) |
| **Filenames** |  |  |  |  |  |
| Sensitivity | 100 (100–100) | 98.47 (97.81–99.08) | 99.94 (99.81–100) | 100 (100–100) | 100 (100–100) |
| Specificity | 98.31 (98.22–98.40) | 99.71 (99.56–99.85) | 98.81 (98.45–99.14) | 100 (100–100) | 99.86 (99.79–99.92) |
| FNR (per-token) | 0.00 (0.00–0.00) | 1.53 (0.92–2.19) | 0.06 (0.00–0.19) | 0.00 (0.00–0.00) | 0.00 (0.00–0.00) |
| FPR (per-token) | 1.69 (1.60–1.78) | 0.29 (0.15–0.44) | 1.19 (0.86–1.55) | 0.00 (0.00–0.00) | 0.14 (0.08–0.21) |
| **Date Shift** |  |  |  |  |  |
| Shift Rate | 98.66 (98.41–98.85) | 99.64 (99.42–99.81) | 99.40 (98.80–99.78) | 99.63 (99.41–99.81) | 98.04 (96.96–98.93) |
| Redaction Rate | 1.21 (1.02–1.45) | 0.09 (0.02–0.18) | 0.27 (0.00–0.80) | 0.05 (0.01–0.12) | 1.30 (0.56–2.25) |
| FNR (per-token) | 0.02 (0.01–0.04) | 0.00 (0.00–0.01) | 0.00 (0.00–0.01) | 0.00 (0.00–0.01) | 0.01 (0.00–0.04) |

\* Sensitivity and specificity refer to deidentification sensitivity and deidentification specificity, respectively. Per-token FNR and FPR quantify errors at the character level (text modalities) or spatial bounding-box level (images and PDFs). Each column reflects a different model used as the MLLM for the orchestrator. All values are percentages with 95% bootstrapped CIs. Intervals reported as 100–100% may arise from zero observed failures or rounding and do not establish perfect performance beyond the evaluated data; see Statistical Analysis. CI denotes confidence interval; FNR, false-negative rate; FPR, false-positive rate; and PII, personally identifiable information.

Deidentification specificity was highest for Qwen3.5-397B-A17B-thinking, followed by Kimi-K2.5-thinking and Qwen3-VL-235B-A22B-Thinking. No clear difference in specificity was observed between Kimi-K2.5-thinking and either Qwen3.5-397B-A17B-thinking (Δ = 0.04 pp; 95% CI, -0.78 to 0.86) or Qwen3-VL-235B-A22B-Thinking (Δ = 0.13 pp; 95% CI, -0.68 to 0.96).

Among the five models, GPT-5.2 had the lowest deidentification specificity and GLM-4.6V the second lowest (Δ = -12.28 pp; 95% CI, -13.84 to -10.34).

The selected local model (Qwen3-VL-235B-A22B-Thinking) achieved deidentification sensitivity with lower confidence bounds exceeding 99% across all six identifier-removal modalities on DeID-Core. Notably, it outperformed GPT-5.2 in date shifting, with fewer inadvertent date redactions.

#### Model scale

Across the Qwen3-VL-Instruct (4B–235B) and Qwen3.5 (9B–397B) families, sensitivity generally increased from the smallest models, but gains diminished and were non-monotonic at larger scales (see Supplementary Figs. S7–S10). Pairwise bootstrap analysis showed higher sensitivity for the 32B than for the 8B Qwen3-VL-Instruct variant (Δ = 0.19 pp; 95% CI, 0.08 to 0.30). For Qwen3.5, no clear sensitivity difference was observed between the 9B and 27B variants (Δ = -0.03 pp; 95% CI, -0.10 to 0.04). Estimated sensitivity differences at larger model sizes had confidence intervals spanning zero for Qwen3-VL-Instruct 30B-A3B vs 235B-A22B (Δ = 0.15 pp; 95% CI, -0.12 to 0.38) and Qwen3.5 35B-A3B vs 397B-A17B (Δ = -0.02 pp; 95% CI, -0.07 to 0.03). Date-shifting performance generally improved relative to the smallest models, although not monotonically with model size. Smaller models tended to redact dates instead of preserving them as instructed.

#### Reasoning capability

The Qwen3-VL-235B-A22B-Thinking variant had higher overall deidentification sensitivity than the Instruct variant (Δ = 0.37 pp; 95% CI, 0.20 to 0.56). Enabling reasoning showed no clear difference for Qwen3.5-397B-A17B (Δ = 0.05 pp; 95% CI, -0.04 to 0.18) and lower sensitivity for Kimi-K2.5 (Δ = -0.91 pp; 95% CI, -1.31 to -0.54) (see Supplementary Figs. S11–S13).

#### Architecture

The dense Qwen3-VL-32B-Instruct and the similarly sized mixture-of-experts Qwen3-VL-30B-A3B-Instruct showed no clear difference in deidentification sensitivity (Δ = 0.11 pp; 95% CI, -0.05 to 0.36) or specificity (Δ = -0.03 pp; 95% CI, -0.89 to 0.82) (see Supplementary Figs. S14–S16).

#### Quantization

Comparing unquantized Qwen3-VL-32B-Instruct with its four-bit AWQ variant showed no clear difference in deidentification sensitivity (Δ = -0.05 pp; 95% CI, -0.13 to 0.03) or specificity (Δ = -0.12 pp; 95% CI, -1.00 to 0.76) (see Supplementary Figs. S17–S19).

#### Reproducibility

We evaluated Kimi-K2.5 (no thinking) across 100 repeated runs with identical inputs and settings (see Supplementary Table S4). Across the repeated runs, text deidentification sensitivity was 98.77% (95% CI, 98.11–99.11). Text sensitivity and text FNR had the widest reported confidence intervals, each spanning 1.00 percentage point; all other metrics showed narrower 95% CIs.

### Benchmark Comparison

We compared the Multimodal Anonymizer using Qwen3-VL-235B-A22B-Thinking with scitlab^13^ and Presidio^14^, selected for their broad overlap with the supported modalities, and ORB-HD^15^ based on CenterFace^16^ (Table 3). The Δ values below derive from the pairwise bootstrap comparisons reported in Supplementary Figs. S20–S21.

**Table 3.**
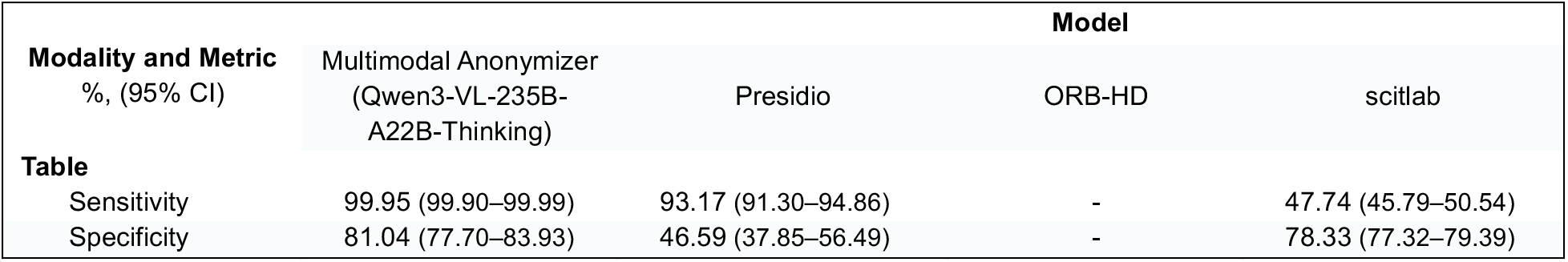
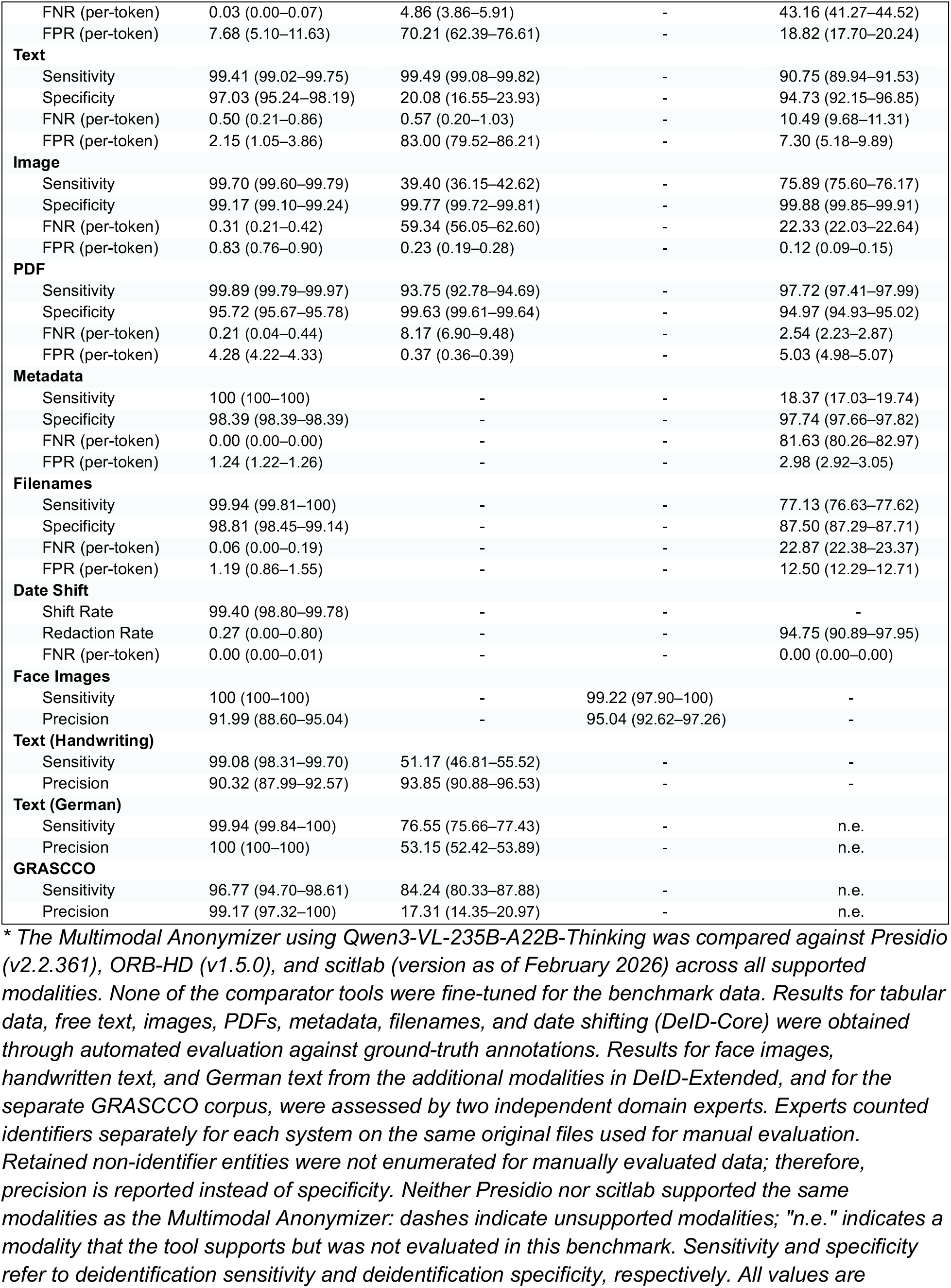
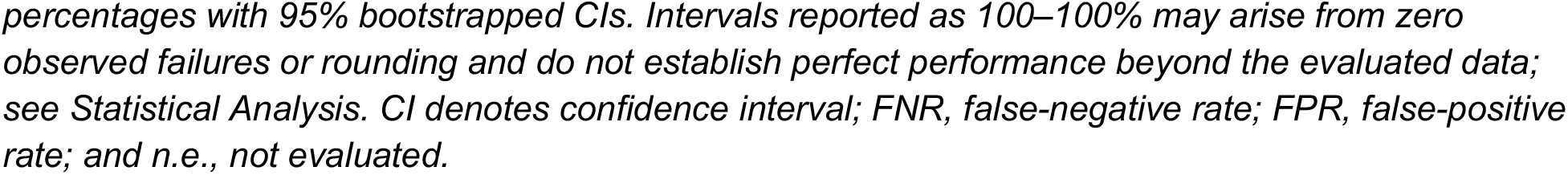
Performance metrics for the benchmark comparison against existing deidentification tools.*

Comparisons used automated evaluation on DeID-Core and expert review of the additional modalities in DeID-Extended and of GRASCCO.

The Multimodal Anonymizer showed higher deidentification sensitivity than Presidio (Δ = 6.78 pp; 95% CI, 5.10 to 8.65) and scitlab (Δ = 52.21 pp; 95% CI, 49.39 to 54.16) for tabular data. Sensitivity was also higher than Presidio (Δ = 60.30 pp; 95% CI, 57.05 to 63.61) and scitlab (Δ = 23.81 pp; 95% CI, 23.51 to 24.12) for imaging data.

On free text, no clear difference in deidentification sensitivity was observed between the Multimodal Anonymizer and Presidio (Δ = -0.07 pp; 95% CI, -0.59 to 0.44), while specificity was higher (Δ = 76.95 pp; 95% CI, 72.72 to 80.89), consistent with greater redaction of non-PII content by Presidio. Compared with scitlab, the Multimodal Anonymizer showed higher text sensitivity (Δ = 8.67 pp; 95% CI, 7.80 to 9.55), with no clear difference in specificity (Δ = 2.30 pp; 95% CI, -0.44 to 5.17).

Metadata and filename deidentification were not implemented in the tested Presidio configuration. Compared with scitlab, the Multimodal Anonymizer showed higher deidentification sensitivity for metadata (Δ = 81.63 pp; 95% CI, 80.19 to 82.98) and filenames (Δ = 22.81 pp; 95% CI, 22.31 to 23.30). Date shifting was not supported by scitlab; instead, it redacted 94.75% (95% CI, 90.89–97.95) of dates.

The Multimodal Anonymizer showed higher deidentification sensitivity than Presidio on handwritten notes (Δ = 47.90 pp; 95% CI, 43.45 to 52.35), German letters (Δ = 23.38 pp; 95% CI, 22.50 to 24.28), and the GRASCCO dataset (Δ = 12.53 pp; 95% CI, 8.35 to 17.08). No clear difference in face detection sensitivity was observed between the Multimodal Anonymizer and ORB-HD (Δ = 0.78 pp; 95% CI, 0.00 to 2.07).

## Discussion

In this study, we developed and evaluated a locally deployable agentic framework for deidentification and observed high rates of identifier removal across the evaluated multimodal clinical datasets, with limited measured loss of clinically important information. The framework showed strong performance across the tested examples of text, tables, PDFs, images, metadata, filenames, audio, and 3D images. It outperformed the tested alternatives in most comparisons and supported a wider range of modalities. These findings provide initial evidence for the feasibility of multimodal deidentification using a locally deployable framework, which, with further validation, could lower a major barrier to the secondary use of hospital data for research and AI development^10,17^.

The importance of these findings is practical as much as technical. In multimodal clinical AI, the central bottleneck is often not model architecture but the inability to safely transform routine hospital data into reusable research assets^18^. Identifiers are distributed across document bodies, headers, overlays, metadata, filenames, and temporal fields, and no single method is likely to be sufficient. Our results are consistent with a systems view of deidentification, in which deterministic rules, specialist models, MLLM reasoning, and verification steps may offer complementary strengths, although their individual and relative contributions require ablation studies.

Consistent with the need to balance disclosure risk and data utility^19–21^, we evaluated deidentification not by sensitivity alone, but jointly with preservation of clinically meaningful information. In clinical records, utility depends not only on retaining individual facts such as diagnoses, medications, and treatment plans, but also on preserving longitudinal temporal structure. In our comparisons, the selected local configuration achieved high identifier removal while limiting unnecessary redaction of clinically important content within the evaluated datasets. Thus, our contribution is the operationalization and empirical assessment of this trade-off for multimodal clinical data deidentification under the evaluated conditions.

Patient-level evaluation was particularly important. In practice, disclosure risk is cumulative: a residual name in one file, an unshifted date in another, and a revealing filename or metadata field elsewhere may together enable reidentification even when document-level performance appears strong. Assessing full multimodal patient records therefore complements token- or document-level metrics alone. The same principle applies to utility, because small errors distributed across multiple files can distort the longitudinal clinical narrative^22^.

These findings also have implications for deployment. The observed performance of a local model is encouraging^10,18^ because health systems often require data residency, auditability, and institutional control. A unified workflow may also facilitate adoption compared with a fragmented collection of modality-specific tools^5,23^. At the same time, date shifting illustrates that deidentification remains a governed trade-off: preserving within-patient temporal structure can improve scientific usefulness, but it may not be appropriate for every data deidentification context and should be paired with setting-specific oversight^24^.

Our study has several limitations: (1) The benchmark relied substantially on MIMIC-IV with synthetic identifier injection, which enabled precise ground truth but may not fully capture the variability of naturally occurring identifiers in routine clinical care. Validation on Charité partograms was limited to one document type at one institution. The observed performance should therefore be interpreted in the context of these datasets, and generalizability across institutions, languages, specialties, and document types warrants further validation. (2) Residual reidentification risk cannot be fully eliminated, particularly through linkage of rare clinical features or external data sources^25–27^. (3) Best performance required substantial computational resources. (4) Data utility was inferred from preservation metrics rather than tested directly in downstream modeling tasks. (5) Component ablations, stronger clinical-text baselines, public-benchmark comparisons, independent security testing, and prospective deployment were not performed.

In summary, our study provides initial evidence that a locally deployable framework can support multimodal clinical data deidentification, with high identifier removal and limited loss of clinically important information in the evaluated datasets. More broadly, the study suggests that deidentification can be operationalized as a unified institutional workflow. With further prospective validation across institutions, systems of this kind could provide valuable infrastructure for the safe reuse of multimodal clinical data in research and AI development.

## Methods

### Study Design and Data Sources

We conducted a retrospective validation study to assess whether a locally deployable agentic system could deidentify multimodal clinical data while preserving clinically important information. We constructed the MIMIC-derived Deidentification Dataset (DeID-Core) using data from 250 patients in the MIMIC-IV database and linked resources, including core hospital and ICU data^28^, emergency department records^29^, clinical notes^30^, chest radiographs^31^, electrocardiograms^32^, and echocardiograms^33^. Because MIMIC-IV is already deidentified, synthetic personally identifiable information was injected into all modalities with exact ground-truth annotations. We then created DeID-Extended by supplementing DeID-Core with publicly available volumetric head CT scans^34,35^, face images^36^, handwritten notes^37^, audio recordings^38^, and synthetic German clinical notes generated by a German physician. For each additional modality, 250 samples were assigned one-to-one to the 250 DeID-Core patient bundles. DeID-Extended therefore includes the complete DeID-Core dataset. These constructed bundles combine unrelated sources and do not represent multimodal records from the same patient; an overview of the datasets used along with further details is provided in Supplementary Figs. S22–S26.

We additionally used the GRASCCO dataset^39^ containing 63 documents as an external German-language text corpus. For site-specific validation, we evaluated the framework on partograms comprising PDF records from 250 distinct patients at Charité – Universitätsmedizin Berlin. The Charité Ethics Committee approved the retrospective use and anonymization of the institution-specific partogram dataset (EA2/267/25) and did not require individual informed consent.

### Benchmark Construction and Ground Truth

Synthetic PII included names, dates of birth, numeric identifiers, and institution-related information. In text and tabular data, synthetic identifiers were inserted into MIMIC-IV-derived content and annotated with exact character positions. In image-based data (ECG PDF files, chest radiograph DICOM images, and echocardiogram DICOM images), synthetic identifiers were rendered with varying fonts, sizes, colors, and positions, and their bounding-box coordinates were recorded as ground truth. Free-text injections were generated with GPT-5.1 assistance and manually verified before evaluation. Additional modalities not amenable to exact automated labeling were assessed by expert manual review.

### Multimodal Anonymizer

We developed the system as a modular framework for multimodal deidentification (Figure 2). Rule-based file-type detection routes each file to a modality-specific processing pipeline, with a central MLLM used where needed. Other deterministic tasks, such as regular-expression-based date detection and shifting, are likewise handled using rule-based methods. Text-based files were analyzed by the MLLM for PII detection and replacement via tool-calling. For images, PDFs, and videos, the pipeline combined optical character recognition with MLLM analysis to identify and redact embedded identifiers. Specialized neural networks were used for facial anonymization in volumetric head scans, face detection in photographs, and handwriting recognition. Audio files were transcribed, deidentified as text, and resynthesized. A verification agent iteratively compared the original and processed outputs to detect residual identifiers or unshifted dates and triggered corrective tools when needed. Outputs preserved the original file format and folder structure. Additional workflow schematics are shown in Supplementary Figs. S27–S28.

**Figure 2:**
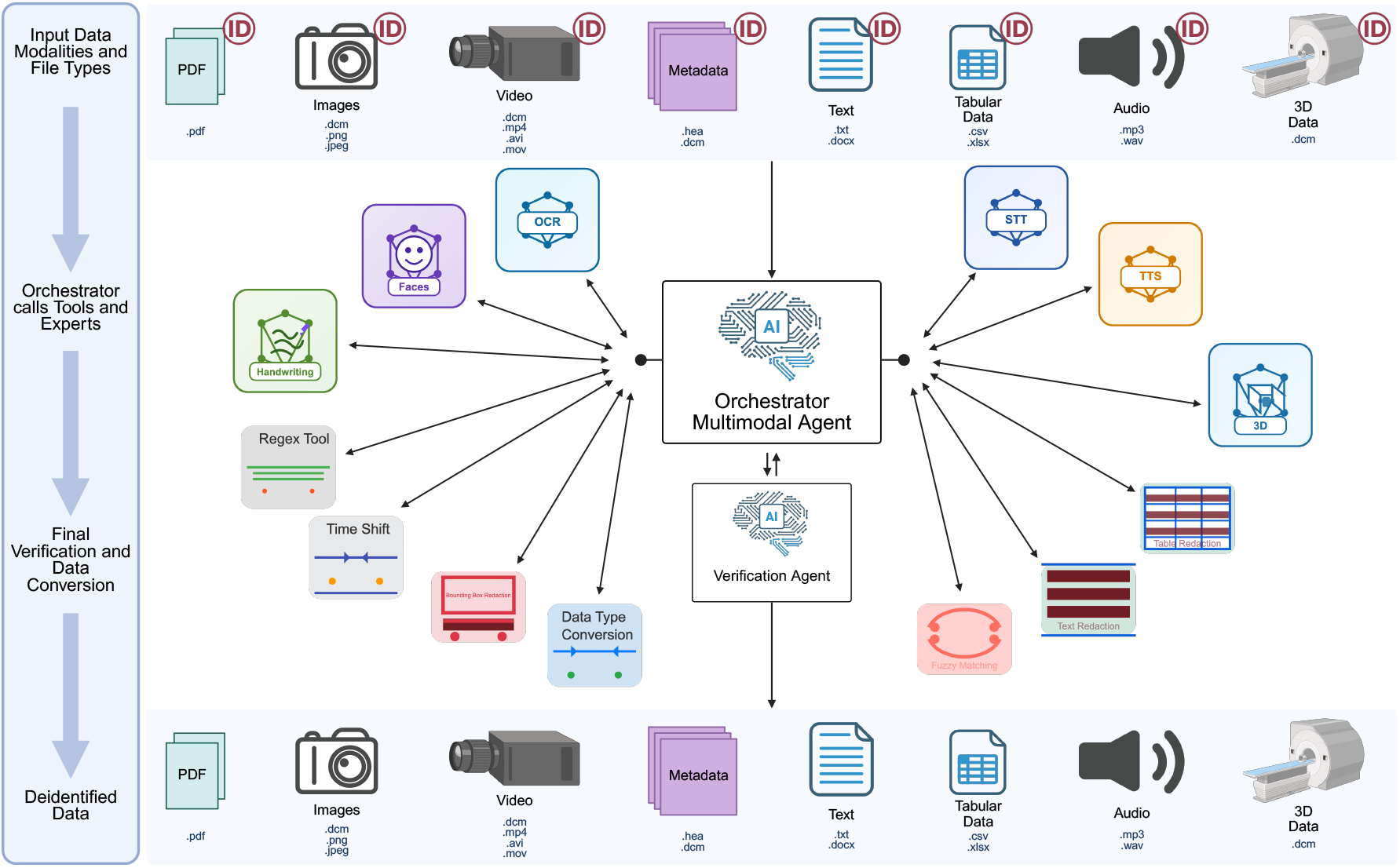
Architecture of the Multimodal Anonymizer. The framework applies modality-specific processing pipelines that are automatically selected based on file-type detection. Additional workflow schematics are provided in Supplementary Figs. S27–S28; here we provide an overview of the key processing steps. The top row shows supported input data modalities and file types, including PDFs, images, videos, metadata, tabular and text-based documents, audio files, and 3D imaging data. As an initial preprocessing step, file and folder names are redacted by the language model, and a patient-specific random time shift (within ± 3 years) is generated and deterministically applied to all dates detected by regular expressions across text-based files. The regex layer covers predefined date formats; non-standard or ambiguous formats missed by regex are handled in the verification step. File-type detection then routes each input to specialized tools and models. Text-based data undergo direct language model analysis for PII identification and replacement with standardized placeholders. Image-based data and PDFs combine OCR (EasyOCR^40^) with MLLM analysis to detect and redact embedded identifiers, such as patient information on radiographs, using fuzzy matching to account for OCR errors. Specialized neural networks address domain-specific challenges, including facial feature anonymization in neuroimaging data (fine-tuned ResNet-UNet on self-annotated 3D data^34^), face detection in images (fine-tuned RetinaNet on an annotated face detection dataset^36^), and handwriting recognition (fine-tuned TrOCR^41,42^ on an annotated handwriting dataset^37^). The ResNet-UNet was trained on approximately 1,000 examples containing faces with approximately 15,000 echocardiography and chest-radiograph images as negative examples and evaluated on approximately 300 examples; RetinaNet was trained on approximately 1,400 face-containing examples with approximately 15,000 negative echocardiography and chest-radiograph images and evaluated on approximately 3,500 examples; and TrOCR was trained on approximately 93,000 examples and evaluated on approximately 23,000 examples. All specialist-model training data were held out from their respective evaluation datasets. Audio processing integrates speech-to-text transcription (whisper-large-v3^43^) with text-to-speech synthesis (Kokoro-82M via kokoro-onnx^44–46^) to produce deidentified recordings. A verification agent iteratively reviews all outputs for residual identifiers or unshifted dates and triggers corrective processing when needed. All pipelines preserve the original file format and return anonymized outputs in the same structure as the inputs (bottom row). CT denotes computed tomography; DICOM, Digital Imaging and Communications in Medicine; MLLM, multimodal large language model; OCR, optical character recognition; PII, personally identifiable information; PDF, portable document format; regex, regular expression; STT, speech-to-text; and TTS, text-to-speech.

### Deidentification Standard

The framework used identifier targets informed by the 18 HIPAA Safe Harbor identifier categories^24^. To preserve longitudinal structure, dates were not removed but replaced by a patient-specific consistent random shift applied across that patient’s records. Date handling was evaluated separately from identifier removal.

### Model Selection and Comparative Evaluation

We evaluated 16 orchestrator model configurations from five families: Kimi-K2.5^47,48^, GLM-4.6V^49,50^, Qwen3-VL^51–58^, Qwen3.5^59–63^, and GPT-5.2^64^. Open-weight models were deployed locally on a cluster with 16 NVIDIA H200 GPUs; GPT-5.2 was accessed through a secured Microsoft Azure deployment as an upper-bound comparator. All models were run with the most deterministic settings available; version details are reported in Supplementary Table S5. We compared model size, reasoning mode, architecture, and quantization. Across these comparisons, the remaining pipeline components—preprocessing, prompts, and specialist models—were held constant.

Following automated evaluation on DeID-Core, Qwen3-VL-235B-A22B-Thinking was selected for the detailed manual evaluation on DeID-Extended. Although Qwen3.5-397B-A17B-Thinking showed marginally higher overall deidentification sensitivity, Qwen3-VL-235B-A22B-Thinking showed higher sensitivity on both text and tabular data and was therefore selected for its more balanced performance across modalities.

For benchmark comparison, we evaluated the Multimodal Anonymizer against existing deidentification tools selected to maximize overlap with the modalities supported by our system. These included scitlab’s proprietary multimodal deidentification tool^13^ and the open-source Presidio^14^ framework, both providing broad deidentification capabilities across multiple data types. As Presidio does not provide image-based face anonymization, we also compared our system with ORB-HD^15,16^, a dedicated face-anonymization method, to extend the open-source comparison to this modality. Comparator tools were run without task-specific fine-tuning or other adaptations.

### Outcomes and Evaluation

The primary manual evaluation on DeID-Extended assessed patient-level and entity-level deidentification sensitivity, patient-level critical clinical preservation, and a clinical preservation score based on file-level redaction annotations. Two independent domain experts reviewed all outputs, with disagreements resolved by consensus. Deidentification sensitivity was assessed per-patient, defined as the proportion of patients with no residual direct identifiers or combination of indirect identifiers across files that could enable reidentification (see Supplementary Table S2), and per-PII entity, defined as the proportion of identifier entities adequately redacted across all modalities. Critical clinical preservation was assessed per-patient and defined as the proportion of patients for whom all clinically essential non-PII information relevant to the current case, such as diagnoses, medications, and treatment plans, remained unaltered. The clinical preservation score was calculated from manual file-level annotations of adequate redactions, excessive redactions and clinical deletions, as described under Statistical Analysis.

Automated secondary analyses on DeID-Core quantified entity-level sensitivity and specificity and per-token false-negative rate (FNR) and false-positive rate (FPR). Deidentification specificity was defined as the proportion of non-PII content correctly left unredacted. FNR and FPR used character counts for text-based modalities and spatial counts for images and PDFs. The FNR was the proportion of PII tokens that remained incorrectly unredacted, whereas the FPR was the proportion of non-PII tokens that were incorrectly redacted. Additional outcomes included date-shift performance, reproducibility, and comparative performance against baseline systems.

Automated evaluation was modality-specific. For text, tables, metadata, and filenames, annotated identifier spans were compared with the redacted output; partially redacted identifiers were counted as failures. For image and PDF modalities, deidentification sensitivity was based on the proportion of annotated bounding-box regions successfully obscured. Date entities were evaluated separately as changed, redacted or unchanged.

### Statistical Analysis

Patient-level metrics were calculated as the proportions of patient bundles meeting the respective outcome criteria. Per-PII sensitivity was calculated from pooled identifier counts. Automated DeID-Core metrics were calculated as ratios of summed metric-specific numerators and denominators, within each modality and across the six identifier-removal modalities for overall estimates. Date handling was evaluated separately. For manually evaluated benchmark comparisons, precision was reported instead of specificity because retained non-identifier entities were not enumerated.

The clinical preservation score was calculated from manual file-level annotations as one minus the number of recorded clinical deletions divided by the total number of adequate and excessive redactions, expressed as a percentage. Clinical deletions were recorded as a subset of excessive redactions. The overall score was calculated by pooling these counts across all evaluated files. For modality-specific scores, counts were first aggregated within each patient and modality to calculate patient-level scores, which were then averaged with equal weight across patients.

Uncertainty was estimated using 10,000 nonparametric bootstrap replicates, with 95% confidence intervals defined by the 2.5th and 97.5th percentiles of the bootstrap distributions. When no failures are observed, percentile-bootstrap confidence intervals can collapse to 100–100%. These degenerate intervals reflect the observed sample and do not establish perfect performance in new data. Identical displayed bounds may also result from rounding. For patient-level outcomes, patient bundles were resampled with replacement, retaining all files associated with each patient together. For per-PII and automated analyses, individual files were resampled with replacement, and metrics were recalculated in each replicate. Between-model differences were calculated within each bootstrap replicate; the reported Δ was the mean of these differences, expressed in percentage points, with percentile-based 95% confidence intervals. Secondary comparisons were exploratory and unadjusted for multiple comparisons.

Reproducibility was assessed across 100 repeated runs of one orchestrator configuration with identical inputs and settings. Supplementary Table S4 reports the arithmetic mean of each metric across these runs. Confidence intervals were calculated using the bootstrap procedure described above, with run-level estimates resampled with replacement.

## Platform Accessibility and Operation

The Multimodal Anonymizer is a fully automated deidentification pipeline, publicly available on GitHub at https://github.com/charite-iaim/multimodal-anonymizer. It can run entirely on local infrastructure using local MLLMs. Base prompts for all supported modalities are provided but are customizable to accommodate specific requirements.

Documentation guides users through installation and configuration. The pipeline is executed via terminal commands or a browser-based interface in three steps:

1. Configure the local MLLM endpoint and select the model.
2. Upload files with optional prompt customization per modality and filename deidentification settings, including the option to preserve a linkage file connecting deidentified and original data.
3. Download processed, compressed output with original folder structures preserved. Once configured, this streamlined interface enables clinical and research personnel to perform comprehensive multimodal deidentification without requiring any programming expertise. Further details on how to use the application, along with example results, can be found in Supplementary Figs. S29–S37.

## Manuscript preparation

ChatGPT (OpenAI) was used during manuscript preparation to assist with language editing and checks of consistency and submission requirements. The authors reviewed and verified the resulting revisions and take full responsibility for the final manuscript.

## Data availability

The datasets used in this study comprise third-party datasets and institution-specific clinical records. The following datasets are available through PhysioNet under their respective access conditions: MIMIC-IV v3.1 (https://doi.org/10.13026/kpb9-mt58); MIMIC-IV-ED v2.2 (https://doi.org/10.13026/5ntk-km72); MIMIC-IV-Note: Deidentified Free-Text Clinical Notes v2.2 (https://doi.org/10.13026/1n74-ne17); MIMIC-CXR Database v2.1.0 (https://doi.org/10.13026/4jqj-jw95); MIMIC-IV-ECG: Diagnostic Electrocardiogram Matched Subset v1.0 (https://doi.org/10.13026/4nqg-sb35); and MIMIC-IV-ECHO: Echocardiogram Matched Subset v0.1 (https://doi.org/10.13026/ef48-v217). Access to restricted resources requires completion of the applicable PhysioNet credentialing, training and data-use agreement requirements.

Additional source datasets are available from their respective providers: the RSNA 2019 Intracranial Hemorrhage Detection dataset through Kaggle (https://www.kaggle.com/c/rsna-intracranial-hemorrhage-detection/data); Computed Tomography (CT) of the Brain through Kaggle (https://www.kaggle.com/datasets/trainingdatapro/computed-tomography-ct-of-the-brain); Face-Detection-Dataset through Kaggle (https://www.kaggle.com/datasets/fareselmenshawii/face-detection-dataset); the IAM Handwriting Database through the official IAM database website (https://fki.tic.heia-fr.ch/databases/iam-handwriting-database), subject to registration and its terms of use; the CSTR VCTK Corpus v0.92 through Edinburgh DataShare (https://doi.org/10.7488/DS/2645); and the Graz Synthetic Clinical Text Corpus (GRASCCO) through Zenodo (https://doi.org/10.5281/zenodo.6539131). Use of these datasets is subject to their respective licences and access conditions.

Additional datasets used for illustrative examples in the Supplementary Information are the RSNA Pneumonia Detection Challenge dataset, available through Kaggle (https://www.kaggle.com/competitions/rsna-pneumonia-detection-challenge/data); PTB-XL v1.0.3, available through PhysioNet (https://doi.org/10.13026/kfzx-aw45); and the 3D echocardiography dataset accompanying EchoSlicer, available through the authors’ GitHub repository (https://github.com/echonet/3d-echo/releases/tag/v1.0). These datasets are subject to their respective access and reuse conditions.

The Charité partogram dataset comprises clinical records from 250 patients and is not publicly available because it contains sensitive patient information and is subject to institutional, ethical and legal restrictions. External access to these records is not provided as part of this publication.

The complete DeID-Core and DeID-Extended benchmarks incorporate restricted third-party data and are therefore not publicly redistributed by the authors. Shareable derived materials, including synthetic-PII injection specifications, benchmark annotations where permitted and aggregate evaluation outputs, are available upon reasonable request to Anja Hirsch. Requests will be assessed against the applicable source-data licences, data-use agreements, institutional policies and patient-privacy requirements. MIMIC-derived data remain subject to PhysioNet’s access and sharing requirements.

## Code availability

The source code for the Multimodal Anonymizer, including the application, prompts, and model-configuration files, is publicly available at the project GitHub repository: https://github.com/charite-iaim/multimodal-anonymizer. The version used for the analyses reported in this manuscript is archived as GitHub release v0.1.0 (https://github.com/charite-iaim/multimodal-anonymizer/releases/tag/v0.1.0) and corresponds to commit ea45dd6e2751c8cbcf9d76add09155fe608d346d. Installation instructions and documentation required to reproduce the processing workflow are provided in the repository.

## Funding

This study was supported by the Berlin Institute for the Foundations of Learning and Data (BIFOLD). AH, RCG, AM, and JM received support from BIFOLD during the conduct of the study. TR and MIG were supported by the BIH Digital Clinician Scientist Program of the Berlin Institute of Health at Charité. The funders had no role in the design or conduct of the study; the collection, management, analysis, or interpretation of the data; the preparation, review, or approval of the manuscript; or the decision to submit the manuscript for publication.

## Author Contributions

AH contributed to the study design, defined the research questions, performed AI inference and statistical analysis, and drafted the manuscript. FWT and KSK performed AI inference. RCG, TR, MIG, RE and MS provided methodological guidance and contributed to the formal analysis. PR provided the Charité data used for analysis. FP designed the study, provided methodological supervision and the technical context of the study. AM designed the study, provided methodological supervision and the technical context of the study. JM designed the study, defined the research questions, performed AI inference and statistical analysis, and drafted the manuscript. AM and JM jointly supervised this work. All authors contributed to revising the manuscript prior to submission.

## Competing interests

Julian Madrid owns shares in Particip GmbH. Alexander Meyer is a co-founder and shareholder of Ascléa GmbH (formerly x-cardiac GmbH), which develops artificial intelligence solutions for healthcare. The remaining authors declare no competing interests.

## Supplementary

**Table S1.**
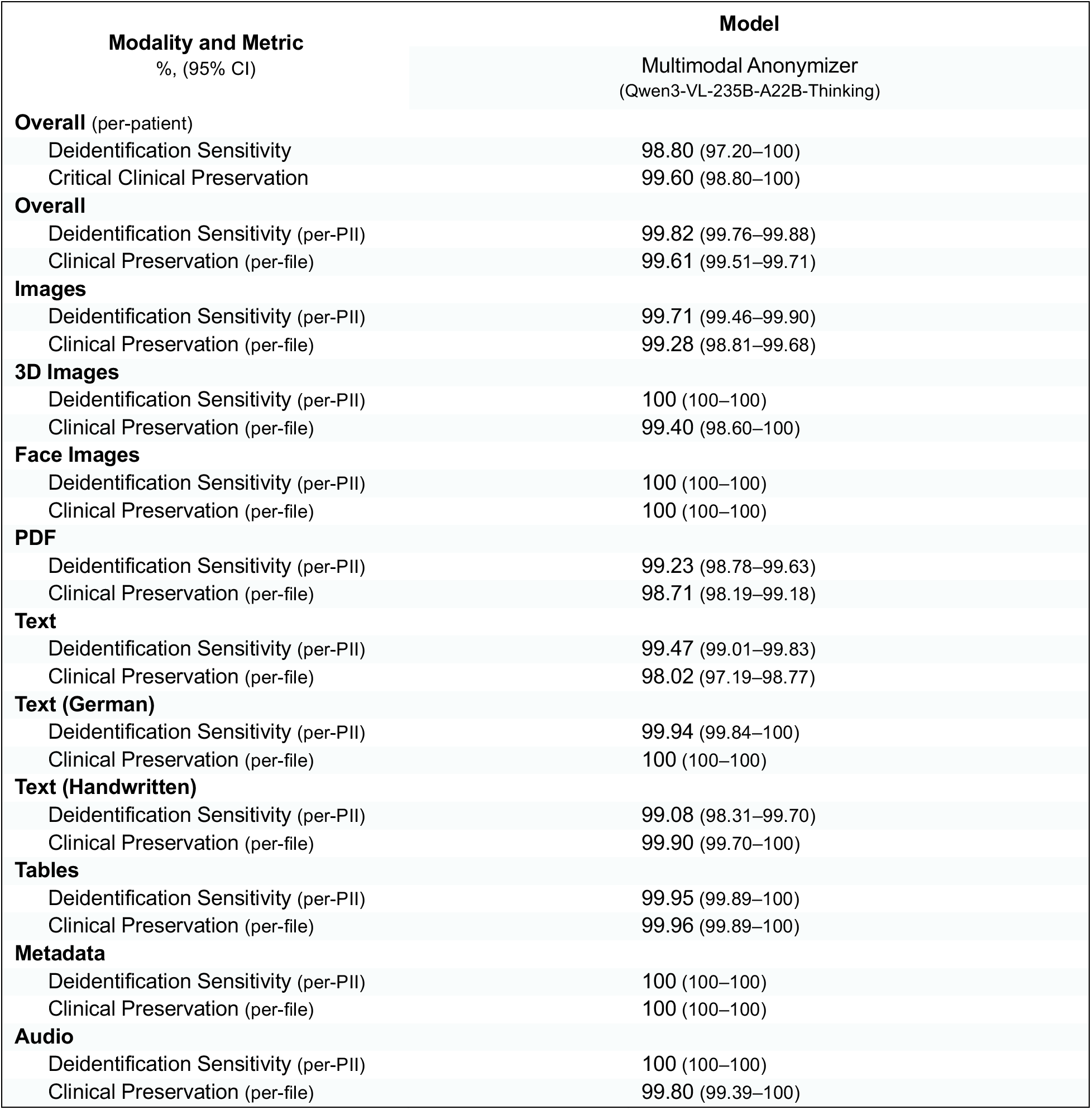
Deidentification Sensitivity, Critical Clinical Preservation and Clinical Preservation Score.

**Table S2.** Manual reidentification assessment criteria.*

| Identifier category | Direct Identifier (one or more identifiers were considered high risk for reidentification) | Indirect Identifier (two or more identifiers were considered high risk for reidentification) |
| --- | --- | --- |
| **Names** | Full name (first name + last name); first name only if uncommon; last name only if uncommon, nickname only if uncommon. | Initials; first name; last name; nickname; name abbreviation; masked name. |
| **Geographic subdivisions smaller than a state** | Full address; GPS coordinates; geocode. | Street name without number; street number without street; neighborhood; city; county; ZIP code. |
| **Dates directly related to an individual** | Age over 89; timestamps with finer than hourly resolution. | Timestamps with hourly or coarser resolution. |
| **Telephone numbers** | Mobile number; work phone; emergency contact number; voicemail number. | Area code; partial phone number. |
| **Fax numbers** | Home fax; employer fax; institutional fax. | Area code; masked fax number. |
| **Email addresses** | Personal email; work email; school email; portal email. | Masked email. |
| **Social Security numbers** | Full social security number; social security number in any format. | Masked social security number. |
| **Medical record numbers** | Medical record number; patient ID; encounter number; accession number if tied to a patient record. | Masked medical record number or patient ID. |
| **Health plan beneficiary numbers** | Insurance member ID; subscriber ID. | Masked member ID or subscriber ID. |
| **Account numbers** | Patient financial account; payment account number. | Masked financial account number or payment account number. |
| **Certificate or license numbers** | Driver's license; passport number; professional license number. | Masked driver's license, passport number or professional license number. |
| **Vehicle identifiers and serial numbers, including license plates** | License plate; vehicle registration number; vehicle serial number. | Masked license plate; masked vehicle registration number; masked vehicle serial number. |
| **Device identifiers and serial numbers** | Medical device serial number. | Masked medical device serial number. |
| **Web URLs** | Patient portal URL; URL containing patient ID; social media account. | Partial URL containing patient ID, masked social media account. |
| **IP addresses** | Static home IP; institutional IP. | Truncated IP. |
| **Biometric identifiers** | Fingerprint; voiceprint; iris scan; retina scan; facial recognition template; palm print; biometric embedding. | Partial biometric identifiers. |

| Table S2. Manual reidentification assessment criteria.* (continued) |  |  |
| --- | --- | --- |
| Identifier category | Direct Identifier (one or more identifiers were considered high risk for reidentification) | Indirect Identifier (two or more identifiers were considered high risk for reidentification) |
| Full-face photographic images and comparable images | Full-face photo; profile face image; video showing face; image where the patient is visually identifiable. | Partial or blurred face, not directly identifiable. |
| Any other unique identifying number, characteristic, or code | Direct identifier if patient is directly identifiable. | Indirect identifier if patient is not directly identifiable. |

*\*We applied a prespecified, rule-based approach to score 250 patient records according to study-specific residual-identifier criteria. Each record was independently reviewed by two expert reviewers for the presence of direct and indirect identifiers. Records were classified as high risk if they contained at least one direct identifier, such as a full patient name or unique identifying number, or at least two indirect identifiers, such as partial identifiers, specific dates, geographic details, or other contextual elements that could enable linkage. Discrepancies between reviewers were resolved by consensus. The direct/indirect distinction was used as an operational re-identification risk framework in this study.<sup>1-4</sup>*

**Table S3.**
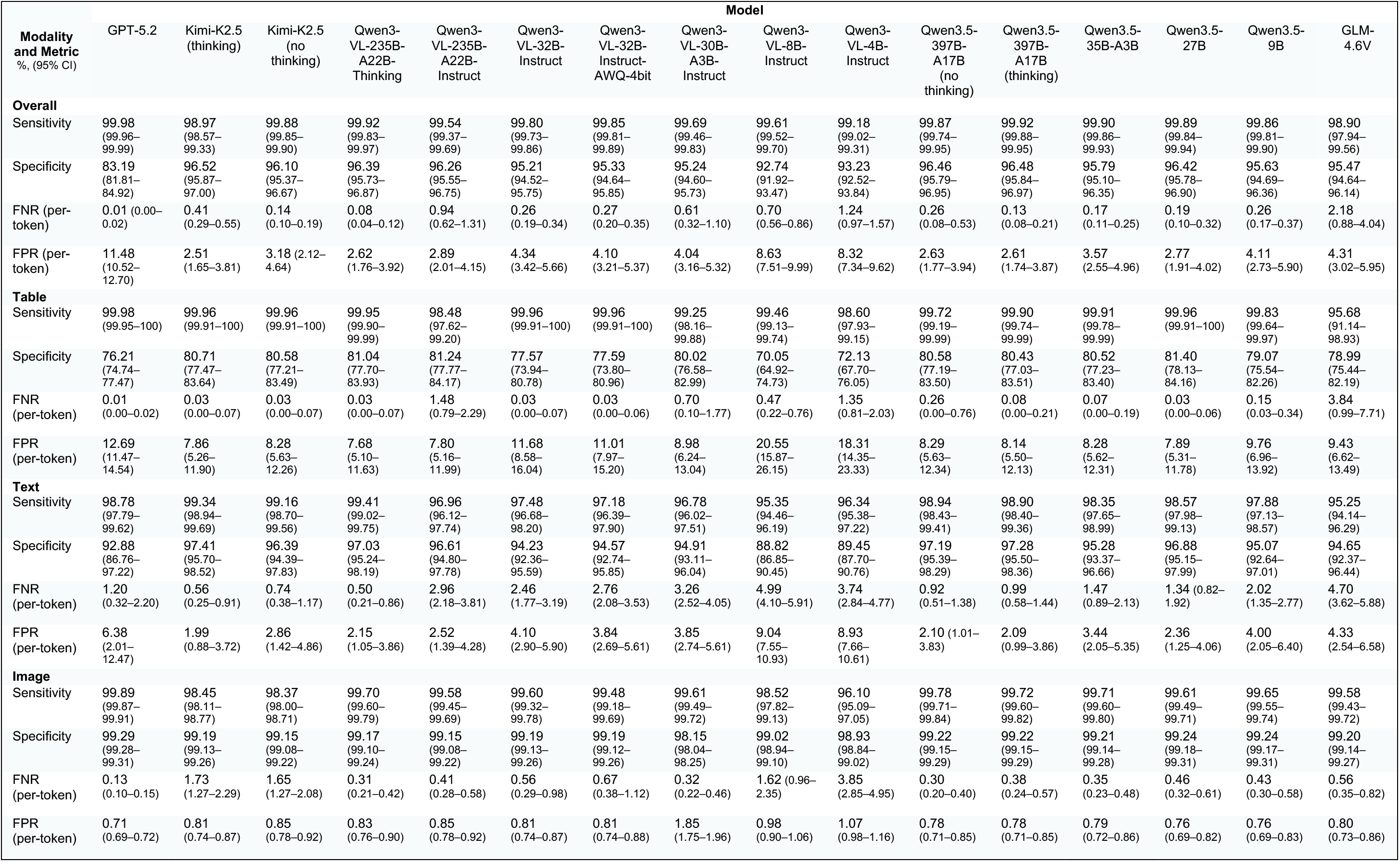
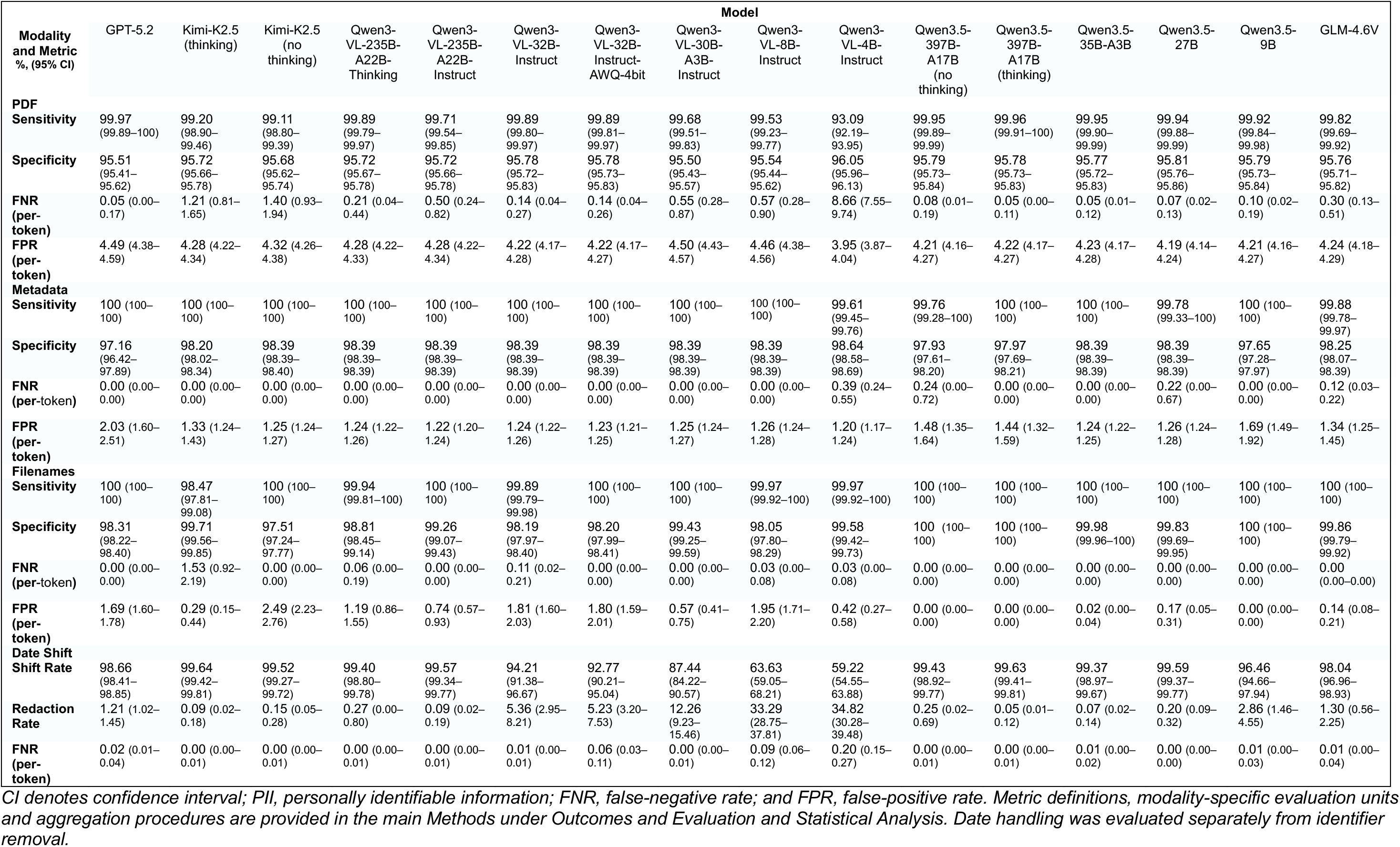
Performance metrics of all tested Models on DeID-Core.*

**Figure S1:**
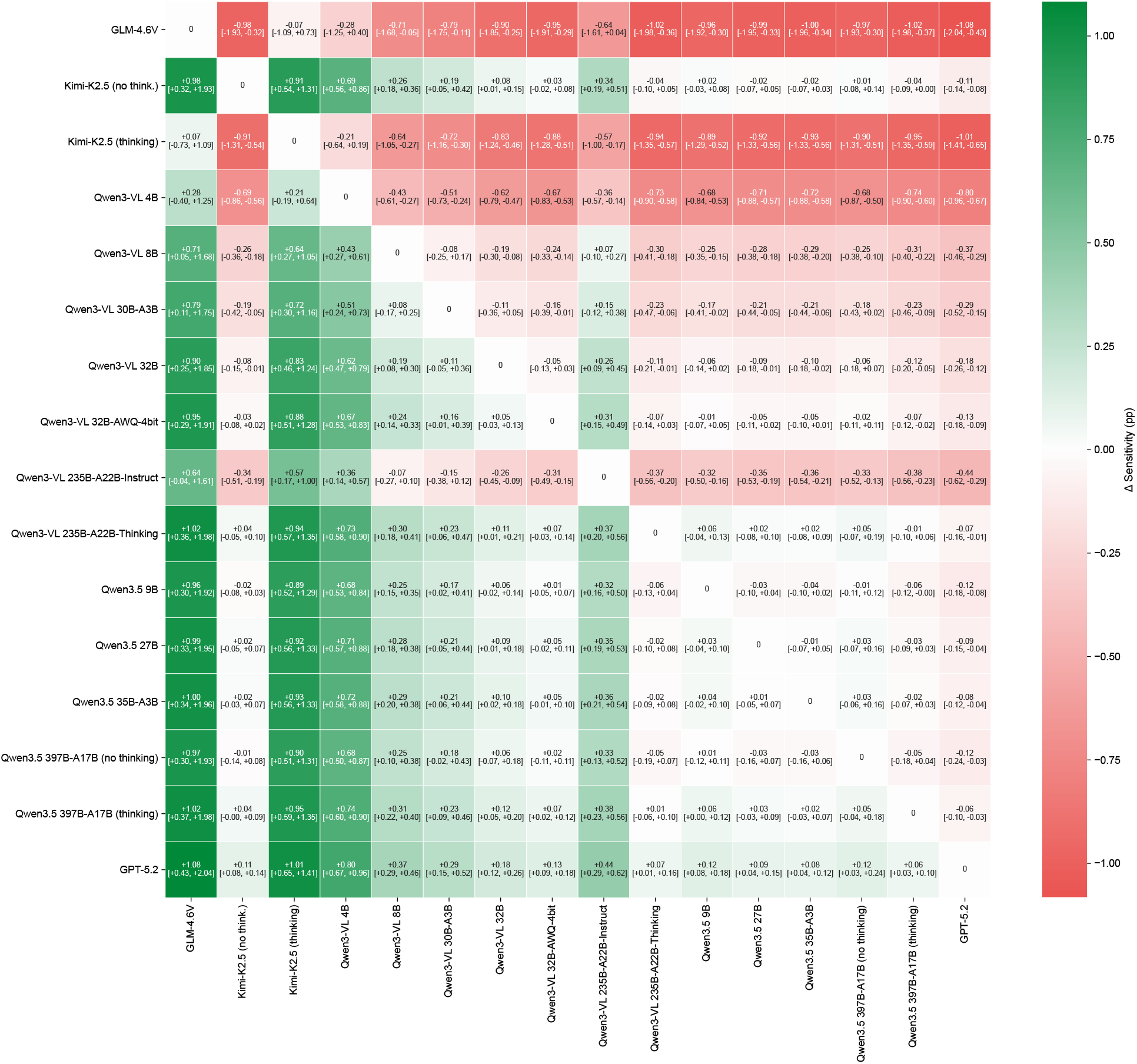
Pairwise bootstrap comparison of deidentification sensitivity (per-PII) across all tested models. Each cell displays the mean difference between the deidentification sensitivities (Δ Sensitivity) of the row model and the column model. Values in brackets denote the 95% confidence intervals. Results were obtained through automated evaluation on DeID-Core.

**Figure S2:**
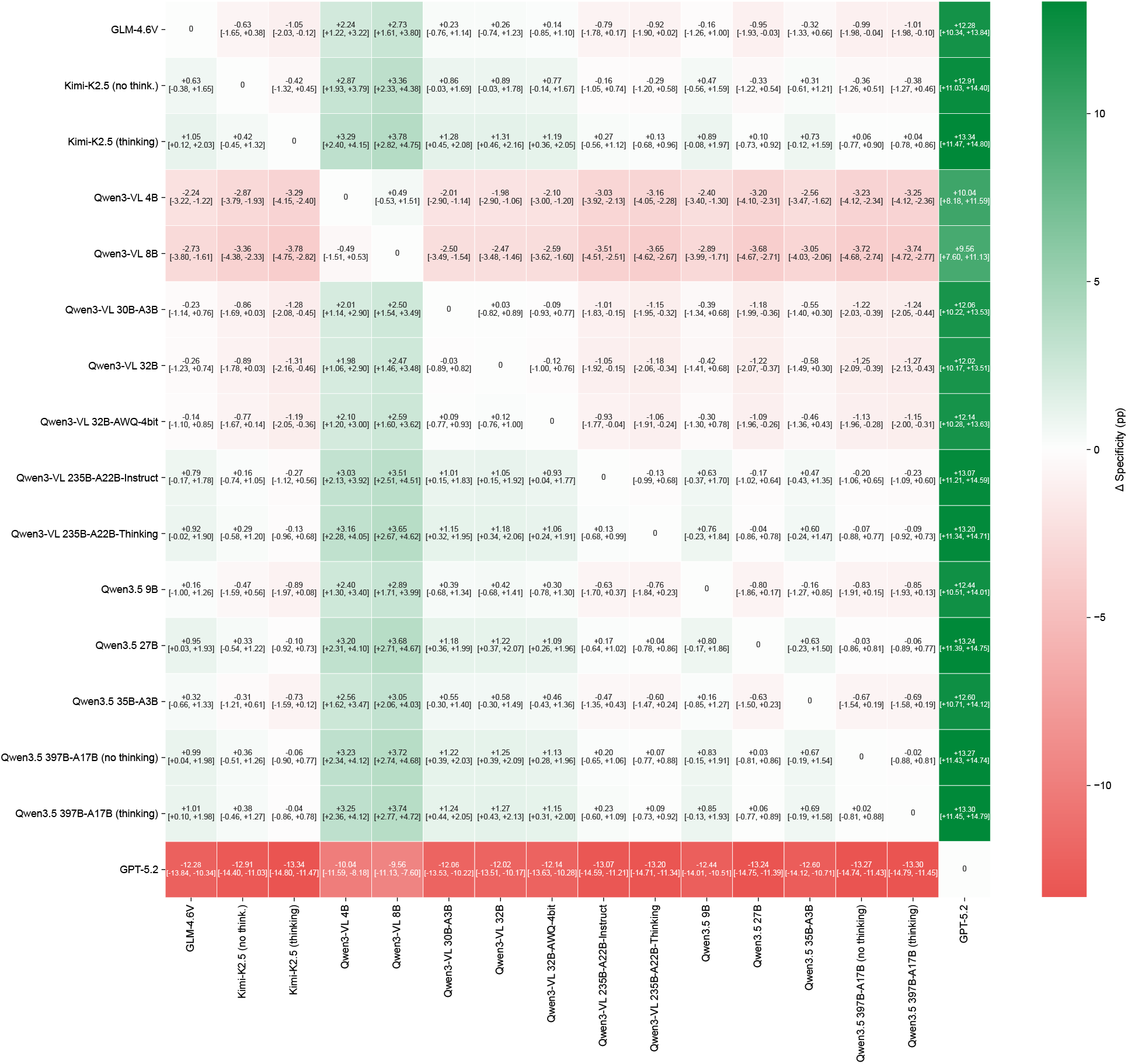
Pairwise bootstrap comparison of deidentification specificity across all tested models. Each cell displays the mean difference between specificities (Δ Specificity) of the row model and the column model. Values in brackets denote the 95% confidence intervals. Results were obtained through automated evaluation on DeID-Core

**Figure S3:**
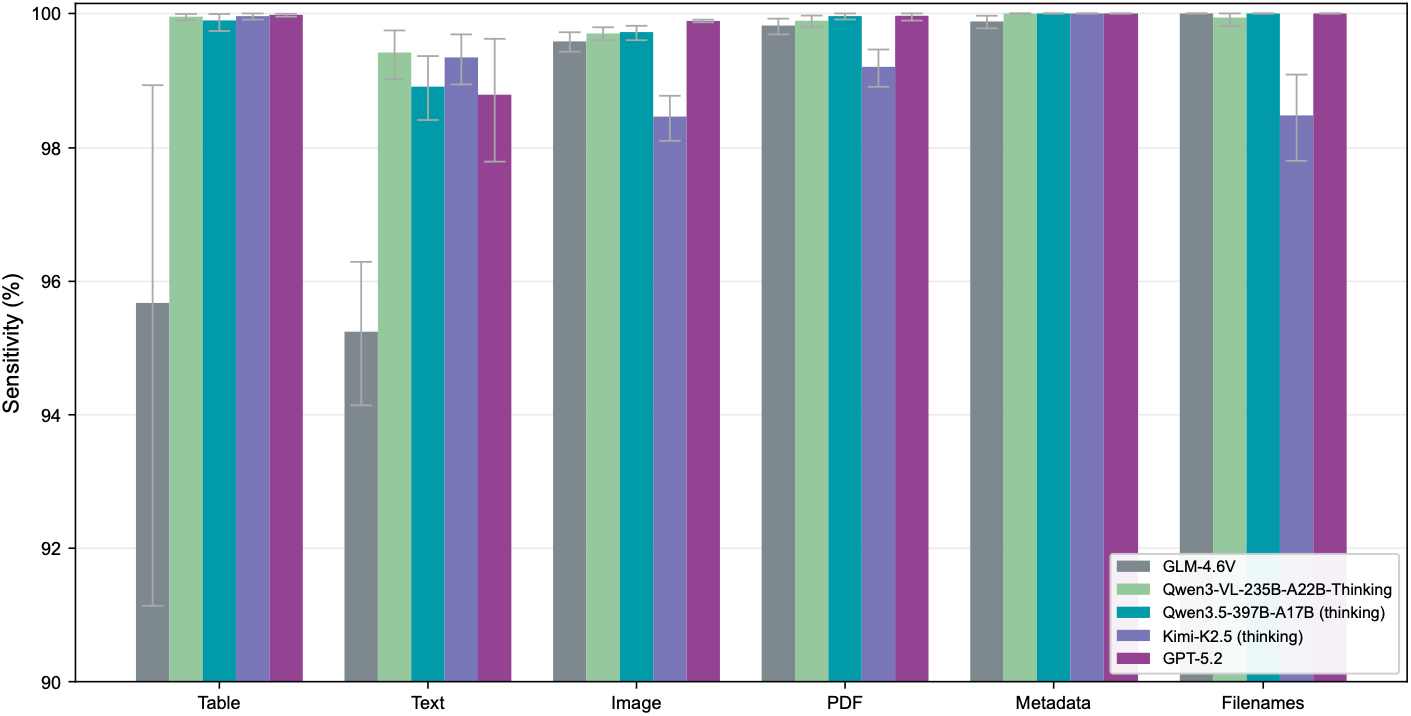
Per-modality deidentification sensitivity (per-PII) of five representative high-capacity models. Bar charts show deidentification sensitivity for the five selected models, each with 95% confidence intervals. Results were obtained through automated evaluation on DeID-Core.

**Figure S4:**
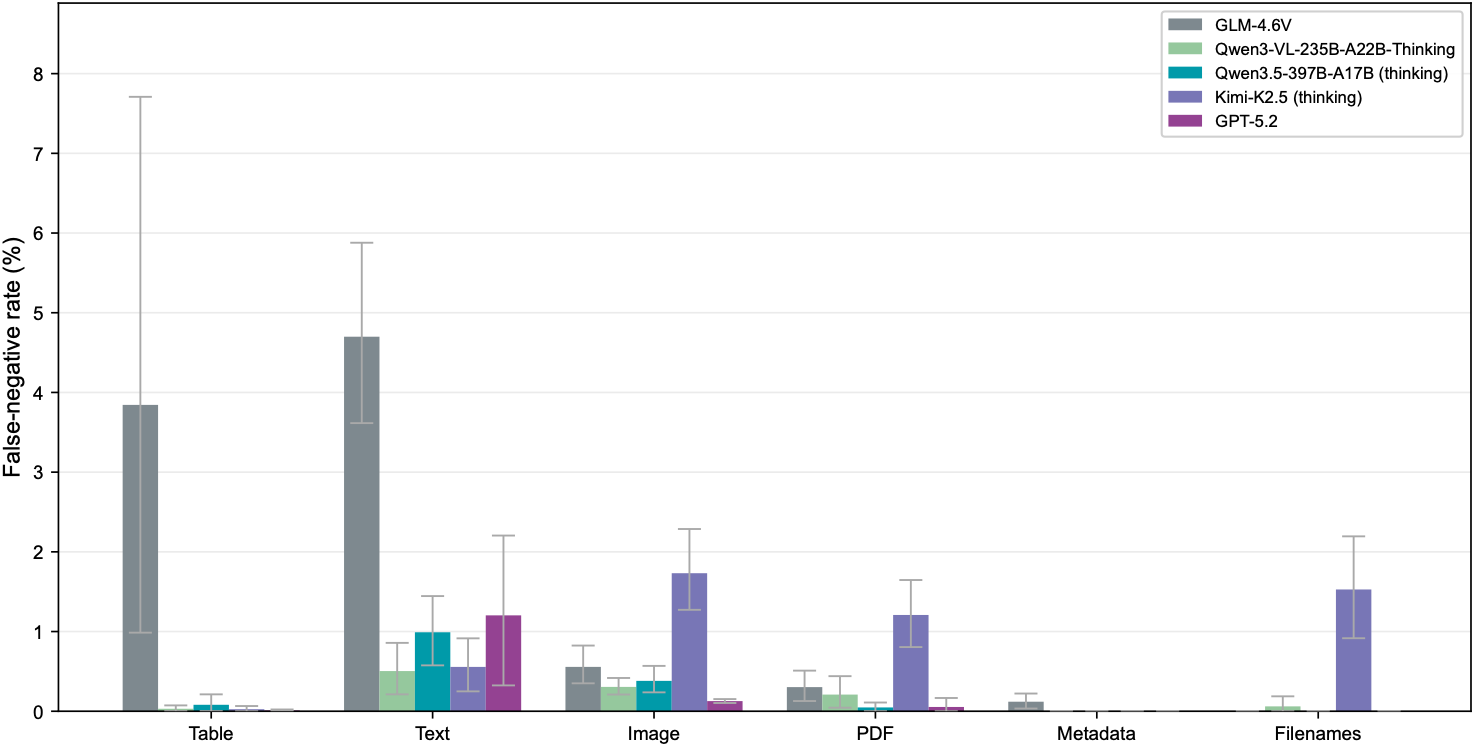
Per-modality deidentification false-negative rates of five representative high-capacity models. Bar charts display the deidentification per-token false-negative rate per modality for the five selected models with 95% confidence intervals. Results were obtained through automated evaluation on DeID-Core.

**Figure S5:**
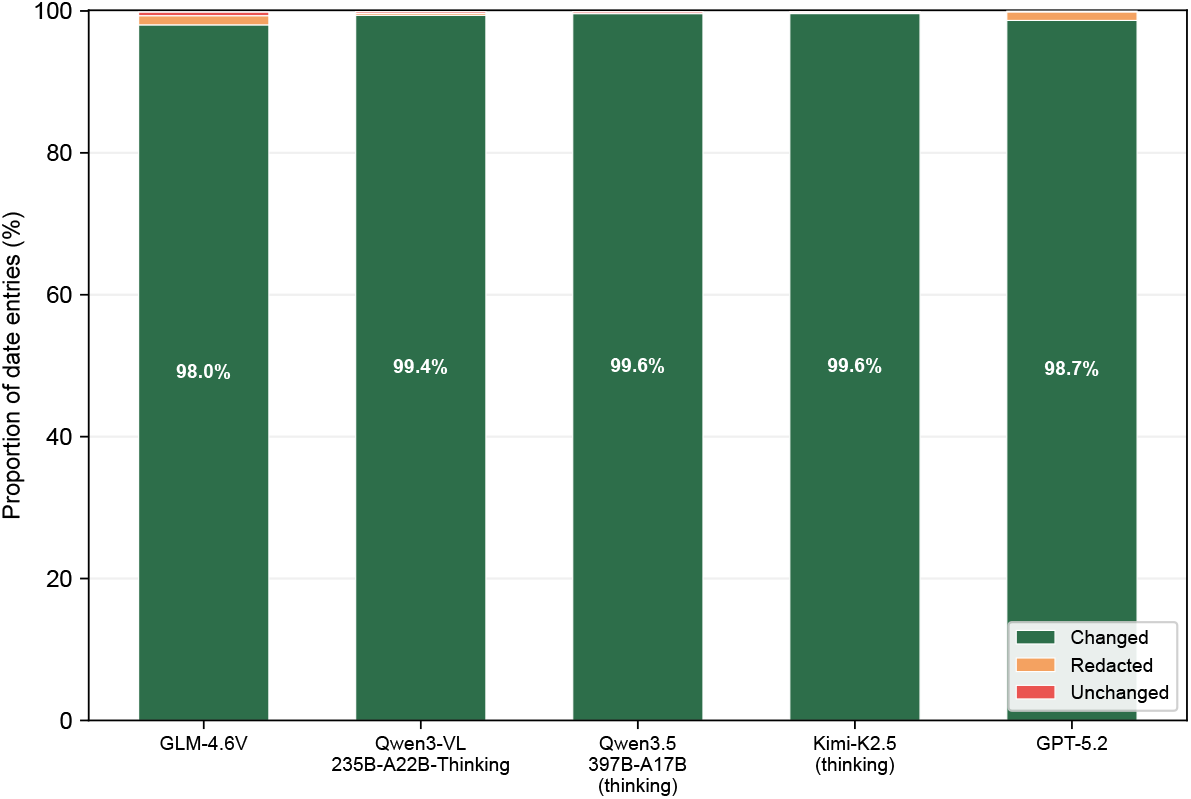
Date shifting performance of five representative high-capacity models. Stacked bar charts display the proportion of date entities that were shifted, redacted, or unchanged across the five selected models. Results were obtained through automated evaluation on DeID-Core.

**Figure S6:**
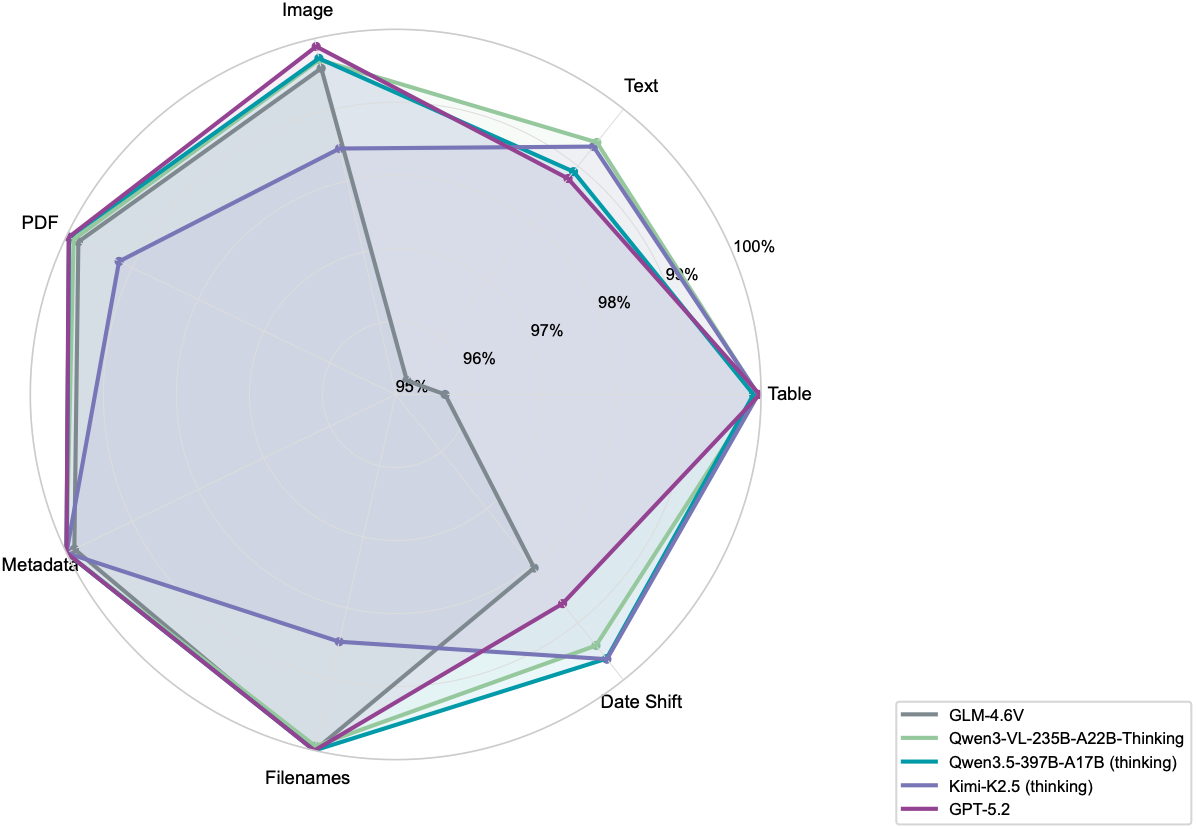
Per-modality deidentification sensitivity profiles of five representative high-capacity models. Per-PII deidentification sensitivity across document element types and date shift (shift rate is displayed) for the five selected models. Results were obtained through automated evaluation on DeID-Core.

**Figure S7:**
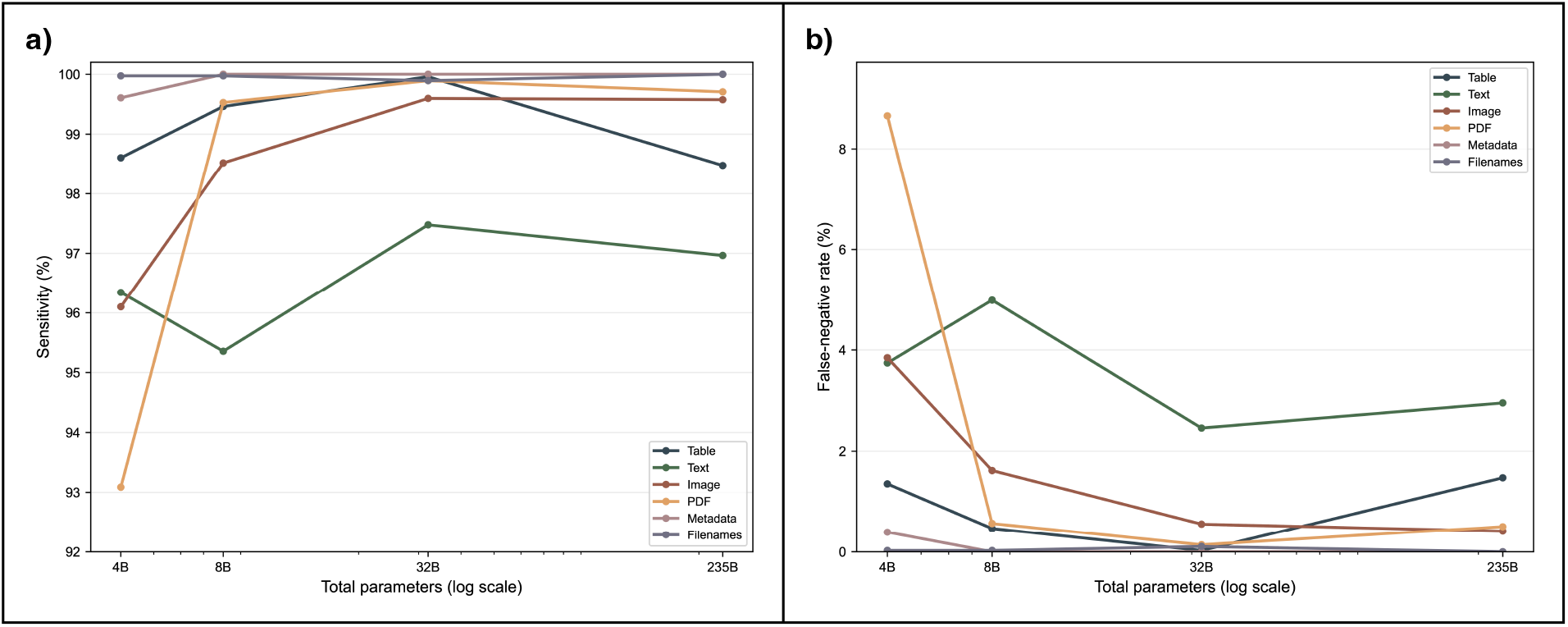
Per-modality deidentification sensitivity and false-negative rate across Qwen3-VL-Instruct model scales. (Panel a) Deidentification sensitivity (per-PII) and (Panel b) false-negative rate (per-token) across Qwen3-VL-Instruct model sizes (4B to 235B parameters), stratified by modality. Results were obtained through automated evaluation on DeID-Core.

**Figure S8:**
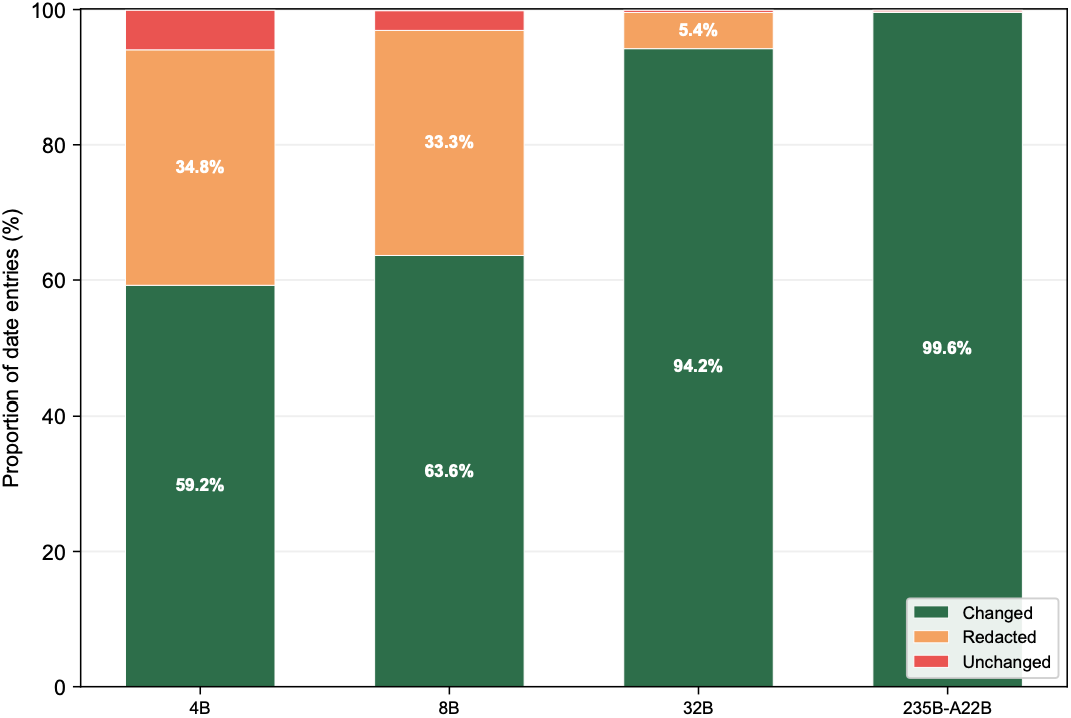
Date shifting performance across Qwen3-VL-Instruct model sizes. Stacked bar charts display the proportion of date entities that were shifted, redacted, or unchanged for four Qwen3-VL-Instruct model sizes (4B to 235B parameters). Results were obtained through automated evaluation on DeID-Core.

**Figure S9:**
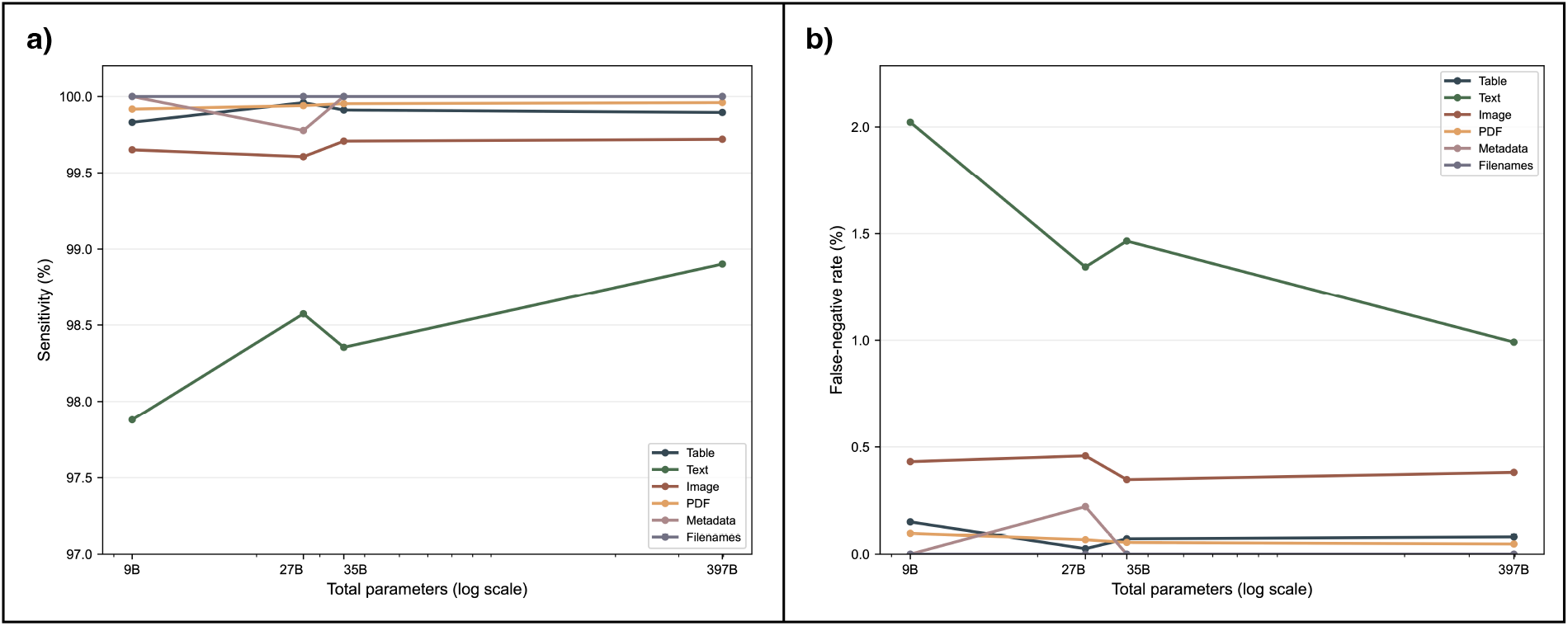
Per-modality deidentification sensitivity and false-negative rate across Qwen3.5 model scales. Per-PII deidentification sensitivity (Panel a) and per-token false-negative rate (FNR) (Panel b) across Qwen3.5 model sizes (9B to 397B-A17B parameters), stratified by modality. Results were obtained through automated evaluation on DeID-Core.

**Figure S10:**
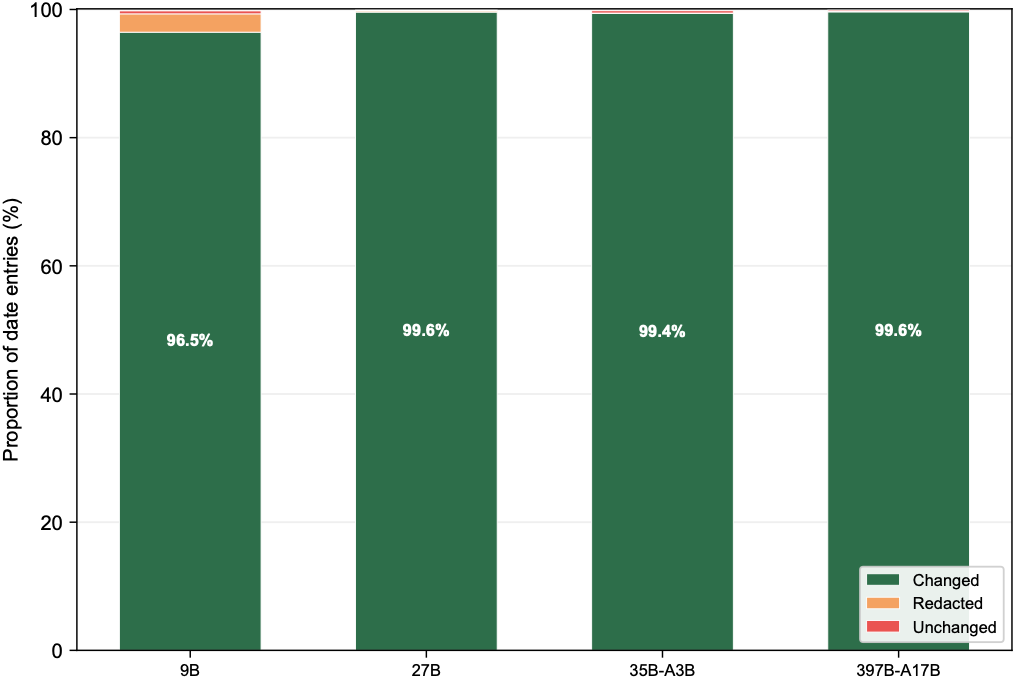
Date shifting performance across Qwen3.5 model sizes. Stacked bar charts display the proportion of date entities that were shifted, redacted, or unchanged for four Qwen3.5 model sizes. Results were obtained through automated evaluation on DeID-Core.

**Figure S11:**
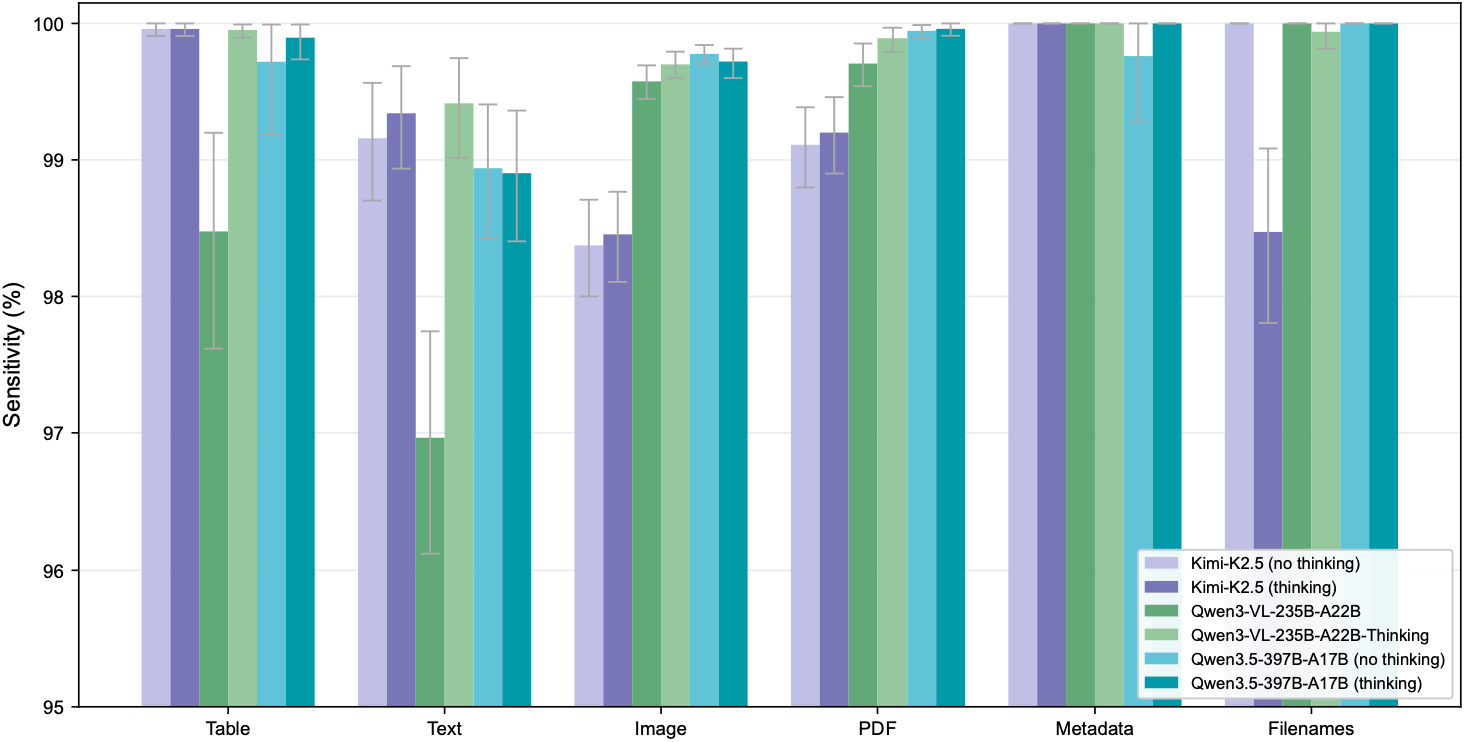
Comparison of thinking modes on per-modality deidentification sensitivity across model variants. Bar charts compare per-PII deidentification sensitivity between thinking and non-thinking variants of Kimi-K2.5, Qwen3-VL-235B-A22B, and Qwen3.5-397B-A17B, with 95% confidence intervals. Results were obtained through automated evaluation on DeID-Core.

**Figure S12:**
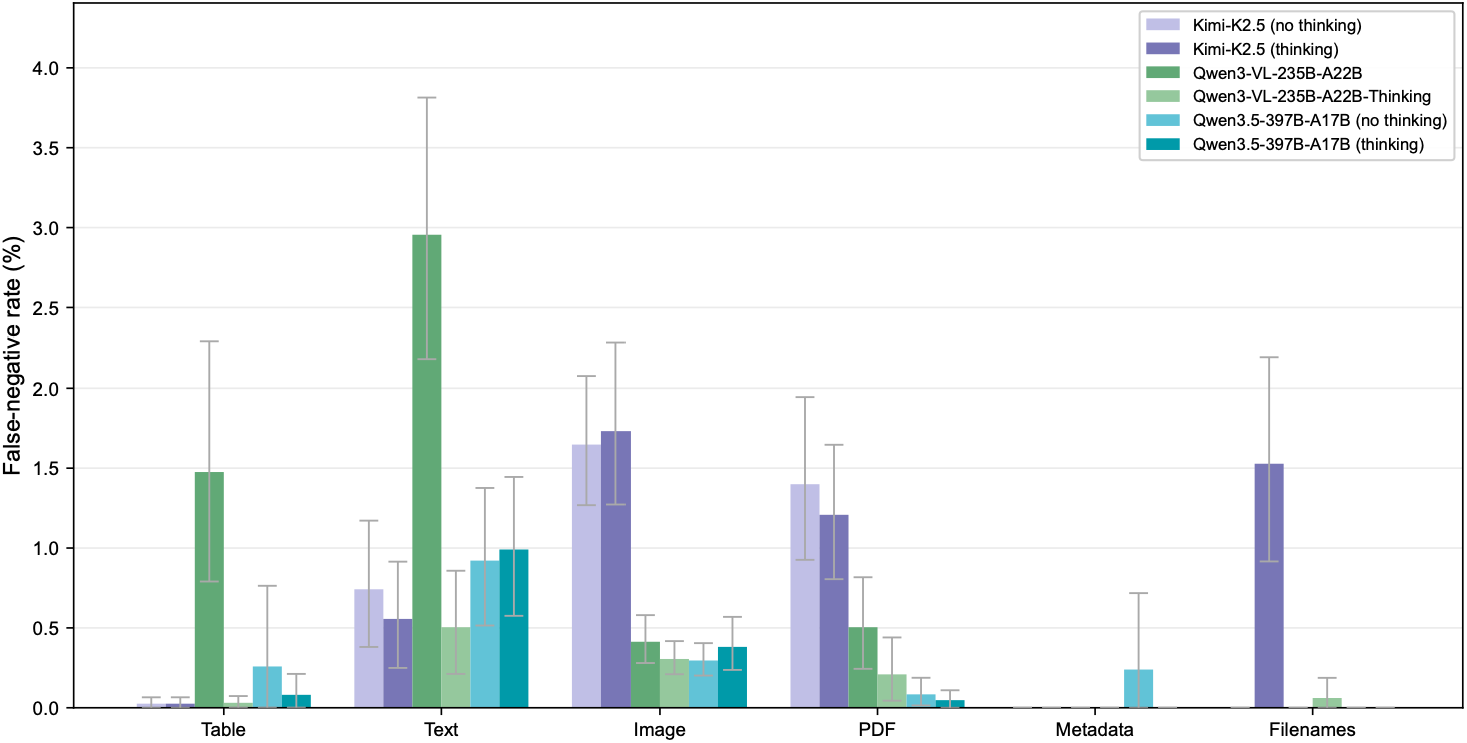
Comparison of thinking modes on per-modality false-negative rates across model variants. Bar charts compare per-token false-negative rates between thinking and non-thinking variants of Kimi-K2.5, Qwen3-VL-235B-A22B, and Qwen3.5-397B-A17B, with 95% confidence intervals. Results were obtained through automated evaluation on DeID-Core.

**Figure S13:**
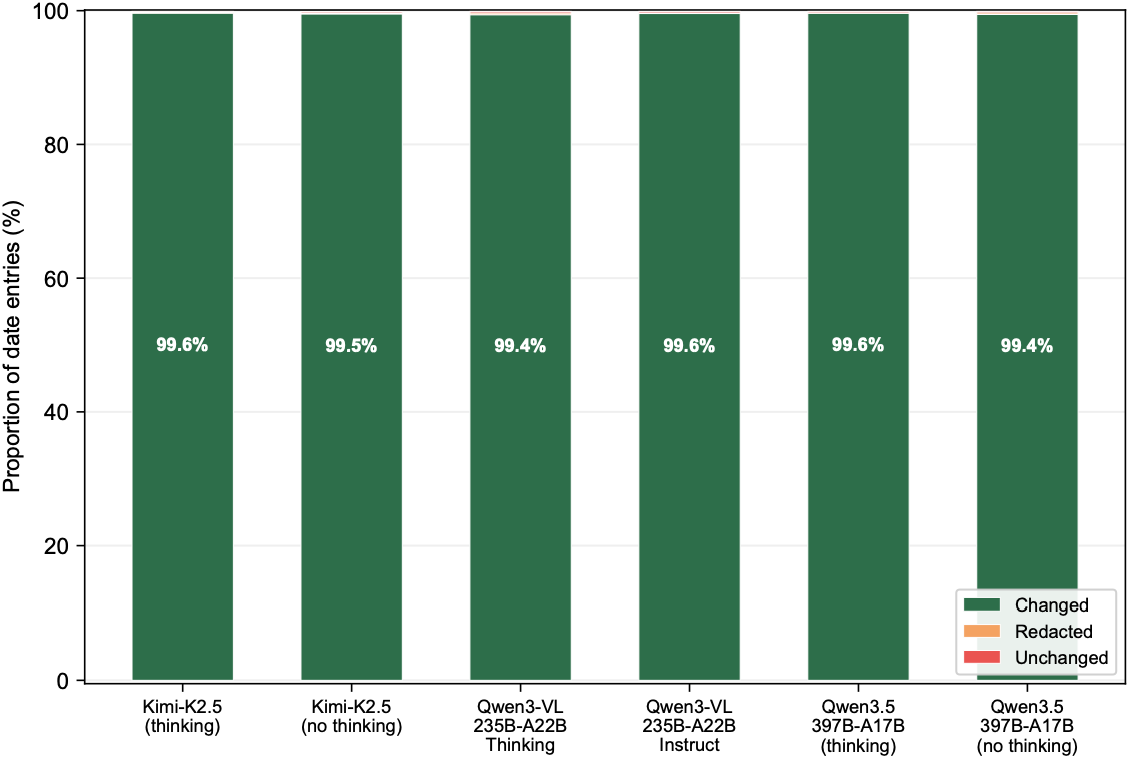
Date shifting performance comparison between thinking and non-thinking model variants. Stacked bar charts display the proportion of date entities that were shifted, redacted, or unchanged for three model families, each evaluated with and without thinking enabled. Results were obtained through automated evaluation on DeID-Core.

**Figure S14:**
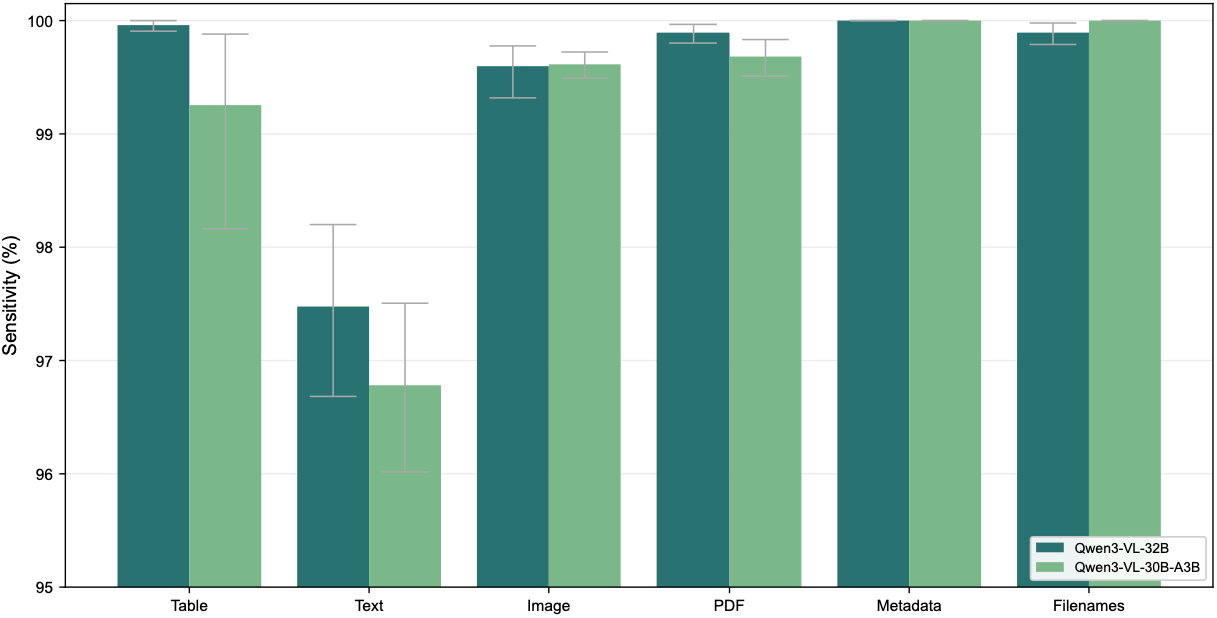
Comparison of per-modality deidentification sensitivity between a dense and a mixture-of-experts model architecture. Bar charts compare per-PII deidentification sensitivity between Qwen3-VL-32B-Instruct (dense) and Qwen3-VL-30B-A3B-Instruct (mixture-of-experts), with 95% confidence intervals. Results were obtained through automated evaluation on DeID-Core.

**Figure S15:**
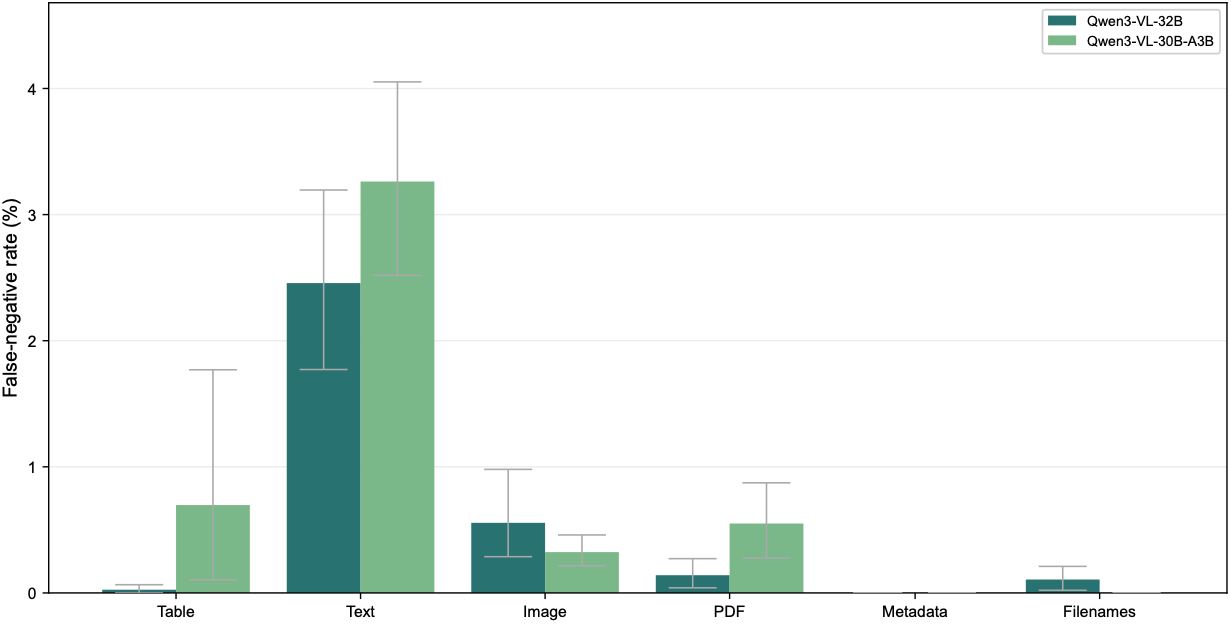
Comparison of per-modality deidentification false-negative rates between a dense and a mixture-of-experts model architecture. Bar charts compare deidentification per-token false-negative rates between Qwen3-VL-32B-Instruct (dense) and Qwen3-VL-30B-A3B-Instruct (mixture-of-experts), with 95% confidence intervals. Results were obtained through automated evaluation on DeID-Core.

**Figure S16:**
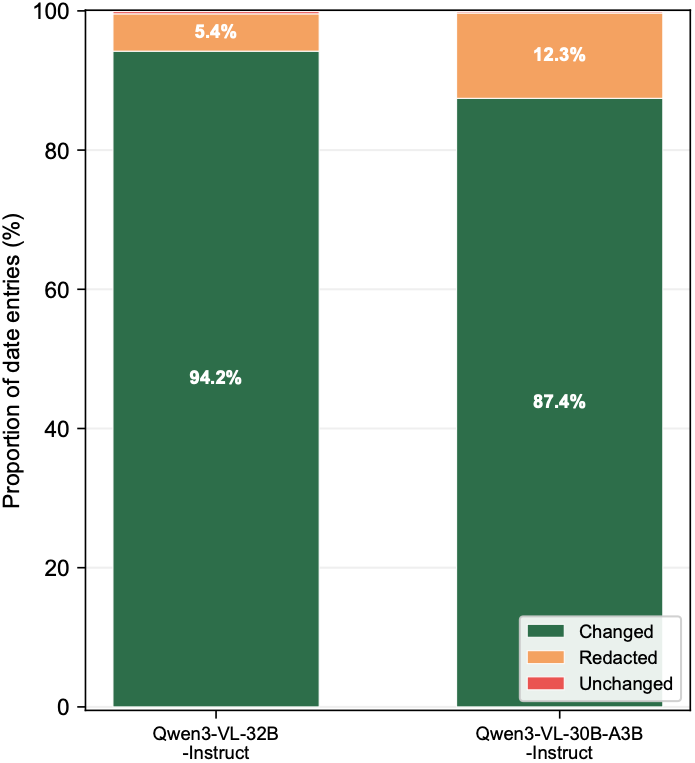
Date shifting performance comparison between dense and mixture-of-experts model architectures. Stacked bar charts display the proportion of date entities that were shifted, redacted, or unchanged for two similarly sized Qwen3-VL-Instruct models with different architectures. Results were obtained through automated evaluation on DeID-Core.

**Figure S17:**
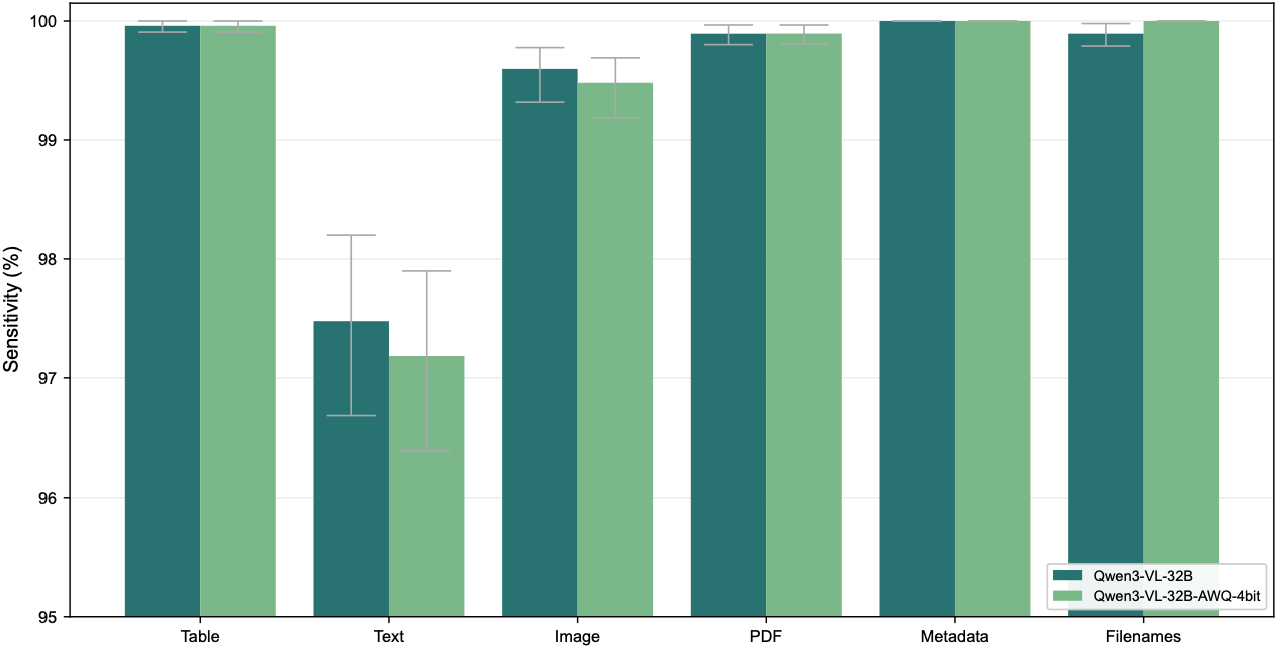
Effect of quantization on per-modality deidentification sensitivity. Bar charts compare per-PII deidentification sensitivity between Qwen3-VL-32B-Instruct (full precision) and its AWQ 4-bit quantized variant, with 95% confidence intervals. Results were obtained through automated evaluation on DeID-Core.

**Figure S18:**
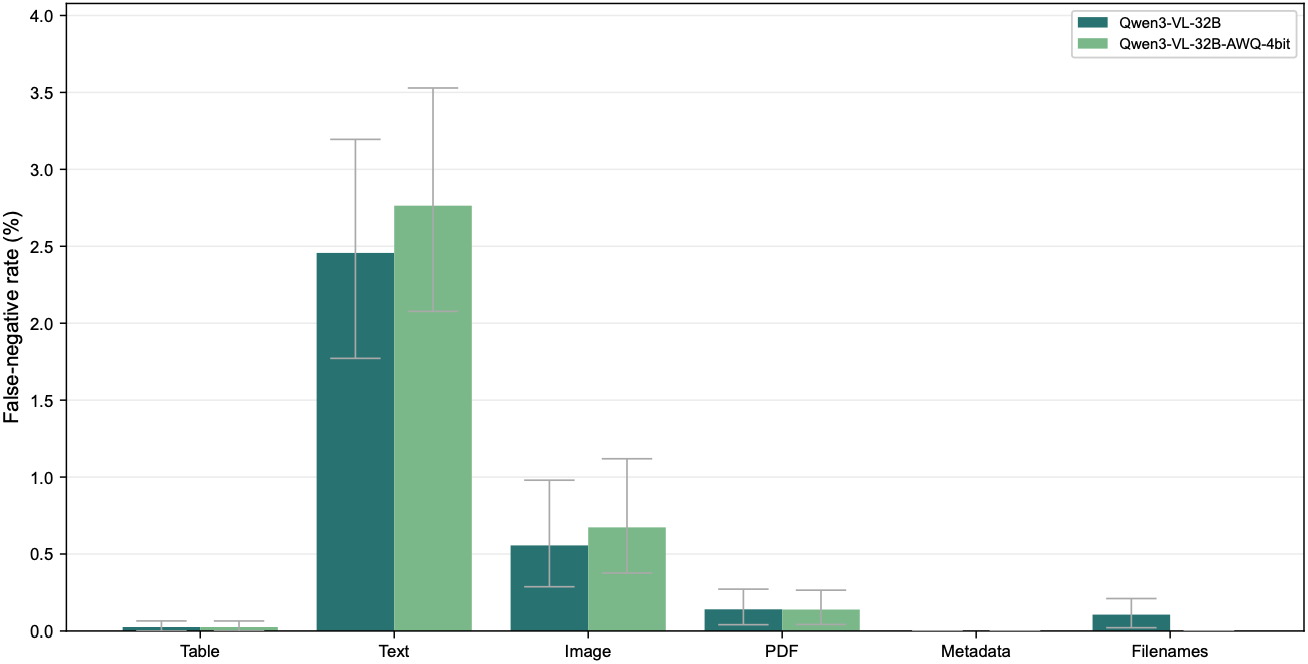
Effect of quantization on per-modality deidentification false-negative rates. Bar charts compare per-token false-negative rates between Qwen3-VL-32B-Instruct (full precision) and its AWQ 4-bit quantized variant, with 95% confidence intervals. Results were obtained through automated evaluation on DeID-Core.

**Figure S19:**
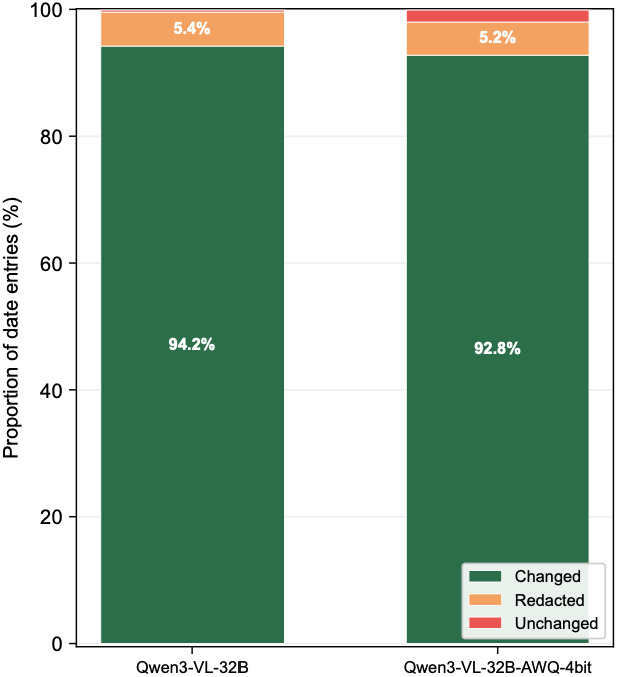
Date shifting performance comparison between quantized and non-quantized model variants. Stacked bar charts display the proportion of date entities that were shifted, redacted, or unchanged for the Qwen3-VL-32B model in its original and AWQ-4bit quantized form. Results were obtained through automated evaluation on DeID-Core.

**Table S4.** Reproducibility Experiment Results on DeID-Core.*

| Table S4. Reproducibility Experiment Results on DeID-Core.* |  |  |  |
| --- | --- | --- | --- |
| Modality and Metric<br>%, (95% CI) | Model | Modality and Metric<br>%, (95% CI) | Model |
| **Table** | Kimi-K2.5 (no thinking) | **PDF** | Kimi-K2.5 (no thinking) |
| Sensitivity | 99.69 (99.16–99.96) | Sensitivity | 99.88 (99.82–99.93) |
| Specificity | 80.74 (80.68–80.79) | Specificity | 95.76 (95.75–95.76) |
| FNR (per-token) | 0.30 (0.03–0.84) | FNR (per-token) | 0.18 (0.13–0.26) |
| FPR (per-token) | 8.11 (8.08–8.13) | FPR (per-token) | 4.24 (4.24–4.25) |
| **Text** |  | **Metadata** |  |
| Sensitivity | 98.77 (98.11–99.11) | Sensitivity | 99.67 (99.02–100) |
| Specificity | 96.50 (96.45–96.55) | Specificity | 98.39 (98.39–98.39) |
| FNR (per-token) | 1.13 (0.79–1.79) | FNR (per-token) | 0.33 (0.00–0.98) |
| FPR (per-token) | 2.74 (2.69–2.79) | FPR (per-token) | 1.24 (1.24–1.24) |
| **Image** |  | **Filenames** |  |
| Sensitivity | 99.63 (99.31–99.81) | Sensitivity | 100 (100–100) |
| Specificity | 99.22 (99.21–99.23) | Specificity | 97.52 (97.51–97.54) |
| FNR (per-token) | 0.44 (0.27–0.74) | FNR (per-token) | 0.00 (0.00–0.00) |
| FPR (per-token) | 0.78 (0.77–0.79) | FPR (per-token) | 2.48 (2.46–2.49) |
|  |  | **Date Shift** |  |
|  |  | Shift Rate | 99.51 (99.48–99.53) |
|  |  | Redaction Rate | 0.16 (0.14–0.19) |
|  |  | FNR (per-token) | 0.00 (0.00–0.00) |

\* Values are arithmetic means across 100 repeated experiments with the same model configuration and data. Sensitivity and specificity refer to deidentification sensitivity and deidentification specificity, respectively. The 95% confidence intervals were calculated using 10,000 bootstrap replicates by resampling run-level estimates with replacement, with bounds defined by the 2.5th and 97.5th percentiles. CI denotes confidence interval; FNR, false-negative rate; and FPR, false-positive rate.

**Figure S20:**
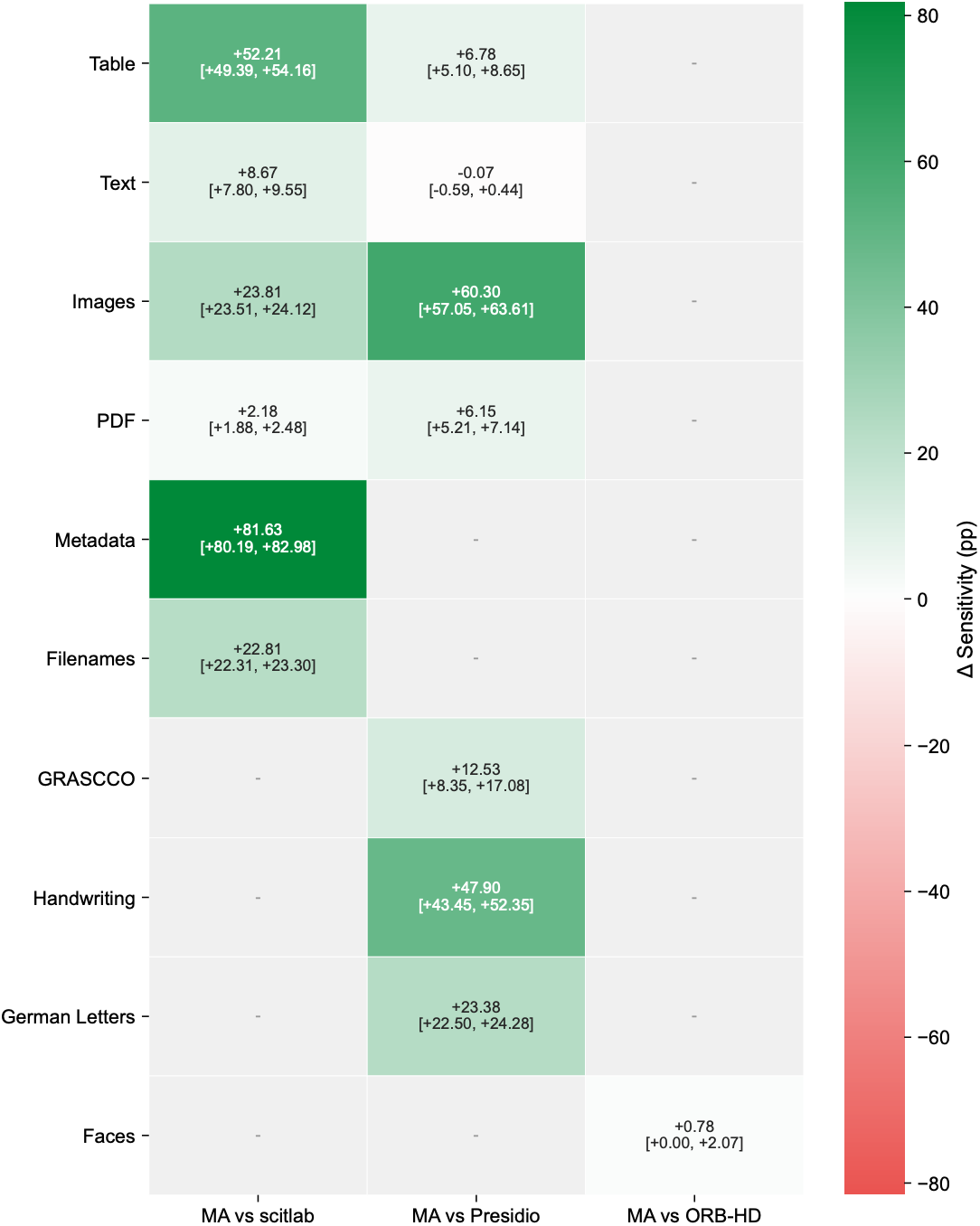
Pairwise bootstrap comparison of deidentification sensitivity per modality between the Multimodal Anonymizer and baseline systems. Each cell displays the mean difference in deidentification sensitivity (Δ Sensitivity) with 95% confidence intervals. Positive values (green) indicate a higher point estimate for the Multimodal Anonymizer (MA) over the respective baseline. Results for tables, text, images, PDFs, metadata, and filenames were obtained through automated evaluation on DeID-Core. Results for handwriting, German letters, and faces from the additional modalities in DeID-Extended, and for the separate GRASCCO corpus, were obtained through expert review.

**Figure S21:**
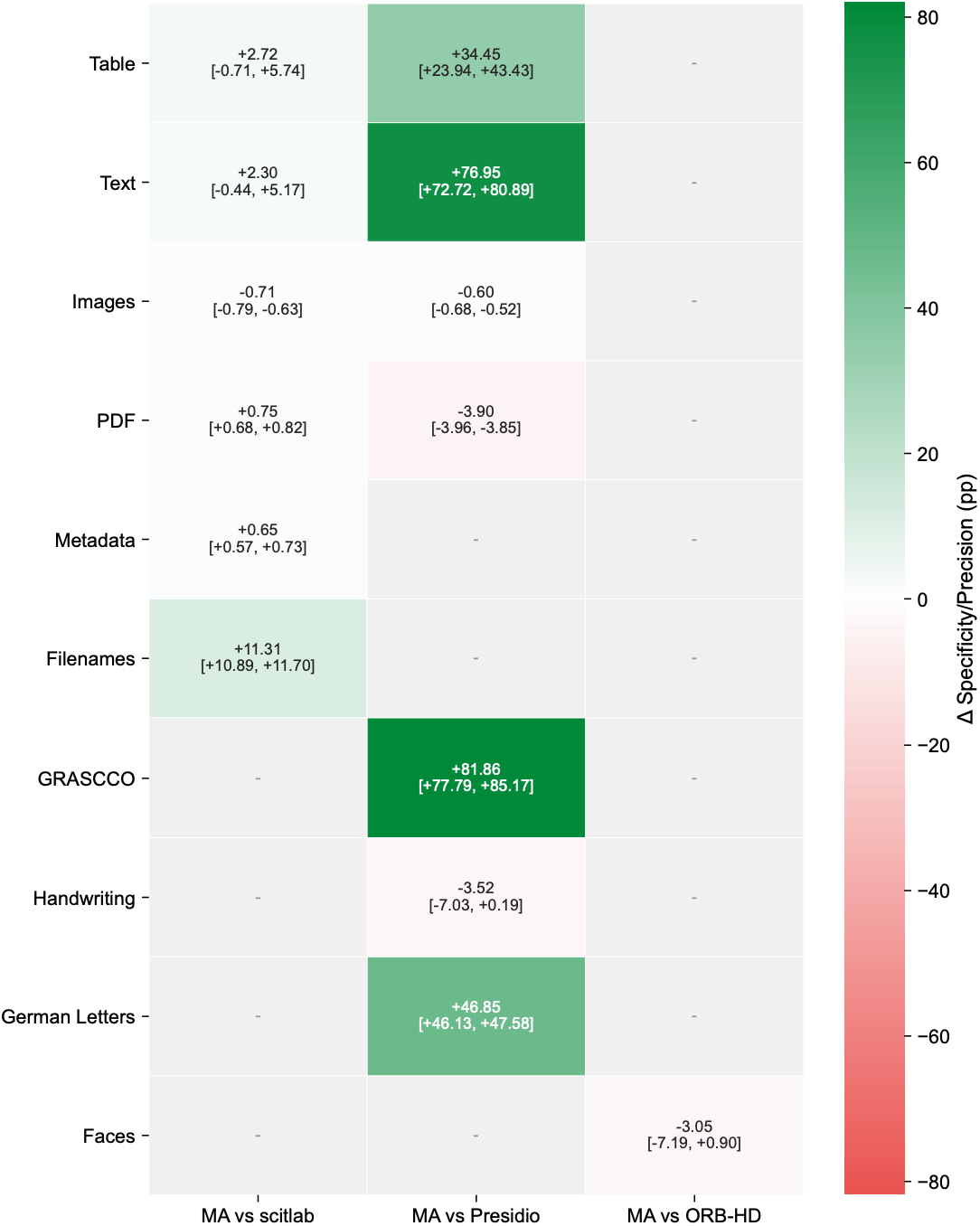
Pairwise bootstrap comparison of deidentification specificity/precision per modality between the Multimodal Anonymizer and baseline systems. Each cell displays the mean difference in deidentification specificity or precision (Δ Specificity/Precision) with 95% confidence intervals. Positive values (green) indicate a higher point estimate for the Multimodal Anonymizer (MA) over the respective baseline. For the automated evaluation on DeID-Core (tables, text, images, PDFs, metadata, and filenames), specificity is reported because true negatives are available. For expert-reviewed handwriting, German letters, and faces from the additional modalities in DeID-Extended, and for GRASCCO, precision is reported because retained non-identifier entities were not enumerated.

**Figure S22:**
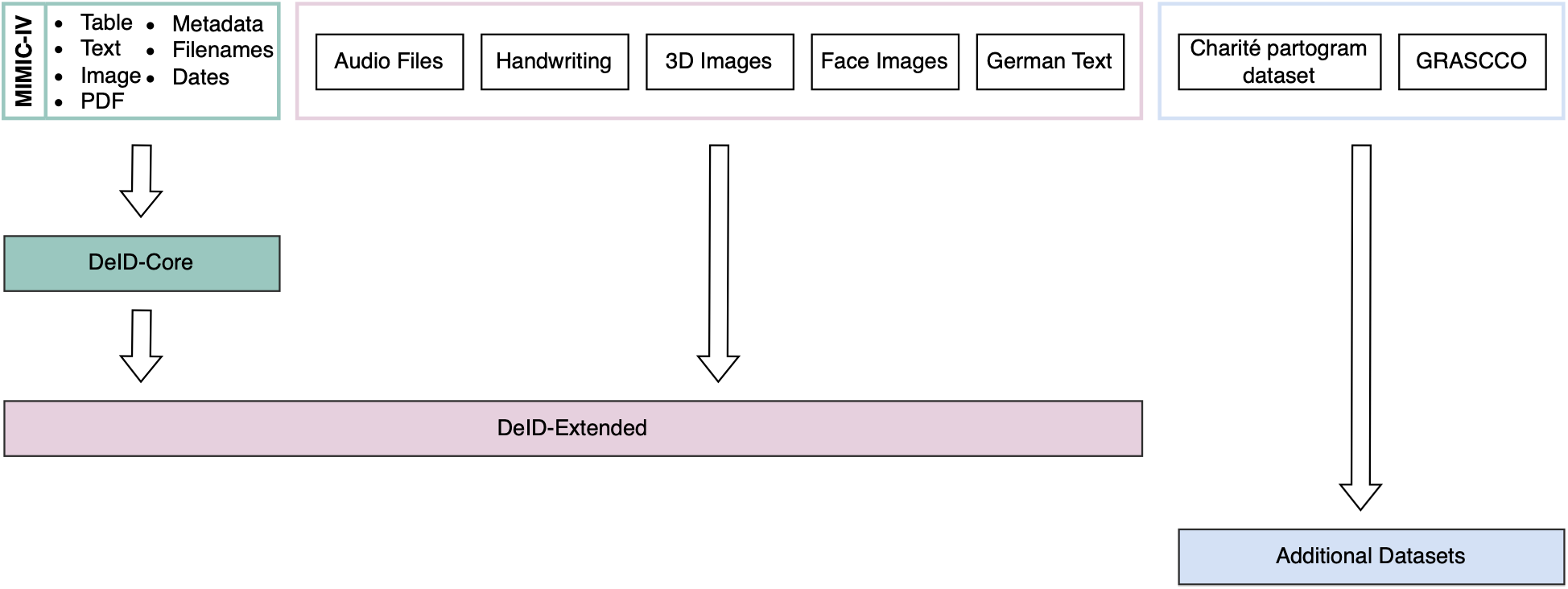
Construction of the Evaluation Datasets. DeID-Core was constructed by injecting synthetic identifiers into deidentified MIMIC data. Additional datasets supplied modality-specific content or identifier surrogates for DeID-Extended; GRASCCO and Charité partograms were evaluated separately. DeID-Core comprises 250 MIMIC-derived patient bundles with injected synthetic identifiers and exact ground-truth annotations. DeID-Extended includes DeID-Core and additional head CT, face, handwriting, audio, and synthetic German-text samples, with one sample per additional modality assigned to each bundle. GRASCCO and the Charité partogram dataset were evaluated separately. During data preparation, electrocardiogram data underwent dual processing. To evaluate performance on the common clinical scenario of scanned or printed ECG reports, we converted the time-series waveform data from its native format into PDF documents. Each PDF was injected with a synthetic patient name, date of birth, and patient identifier, along with additional randomly selected fields. These included further PIIs such as physician name, study date, and institution name, as well as non-PII values such as heart rate, PR interval, and QRS duration. Identifiers were placed at several randomly selected positions within each file to evaluate detection at different locations. ECG header metadata files were annotated separately; their clearly structured format allowed precise specification of identifier locations for subject identifiers and date information. Chest radiographs as well as echocardiogram videos and images in DICOM format received identifier injections analogous to those used for the ECGs. Bounding box coordinates were recorded for all injected identifiers, enabling the PIIs to be localized for subsequent evaluation. Tabular data presented unique challenges due to the complexity and volume of clinical database exports. We derived CSV files from the MIMIC-IV database structure. Annotation followed an XML-style scheme, with personal identifiers marked by <PER> tags and temporal information by <DATE> tags. Columns containing PIIs, such as subject and admission identifiers, were annotated by explicitly defining which columns constitute PIIs. Where appropriate, we added further PII columns containing, among other things, names, social security numbers, and addresses. Free-text columns, including clinical comments, underwent combined annotation and injection using GPT-5.1. Discharge summaries and radiology notes received an enriched injection of identifiers such as names, locations, and telephone numbers, with all generated annotations subsequently verified and supplemented manually. The textual data used for evaluation consisted of the annotated and injected discharge-summary text columns, exported as standalone text files. Finally, file and folder names throughout the dataset were annotated for any embedded identifiers using <PER> tags, allowing us to assess whether personal identifiers in filenames are correctly redacted.

**Figure S23:**
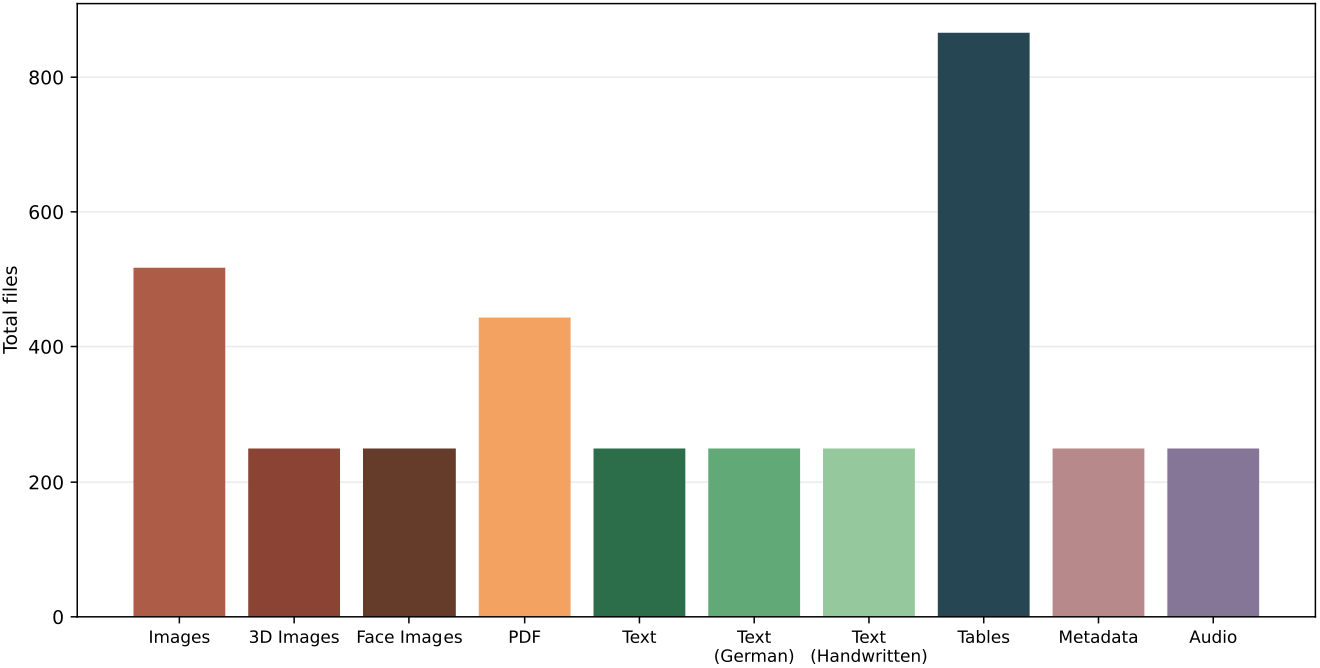
Distribution of files per modality in DeID-Extended.

**Figure S24:**
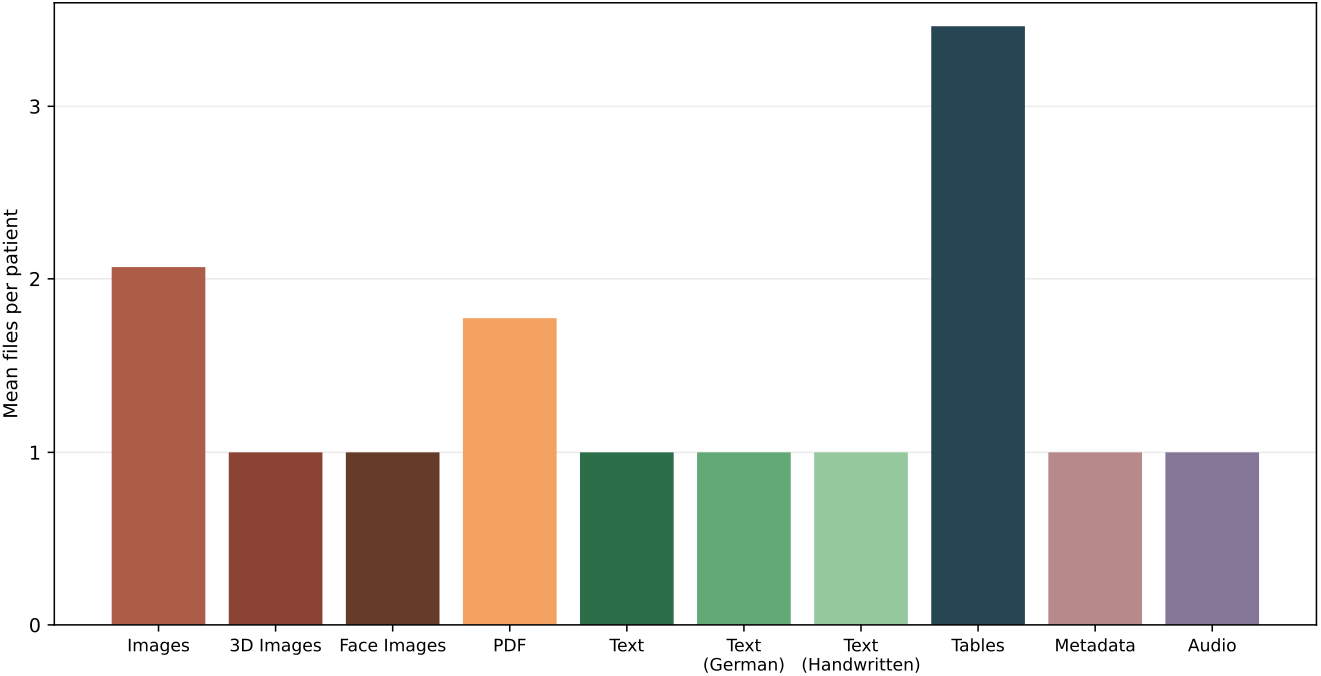
Mean number of files per patient across modalities in DeID-Extended.

**Figure S25:**
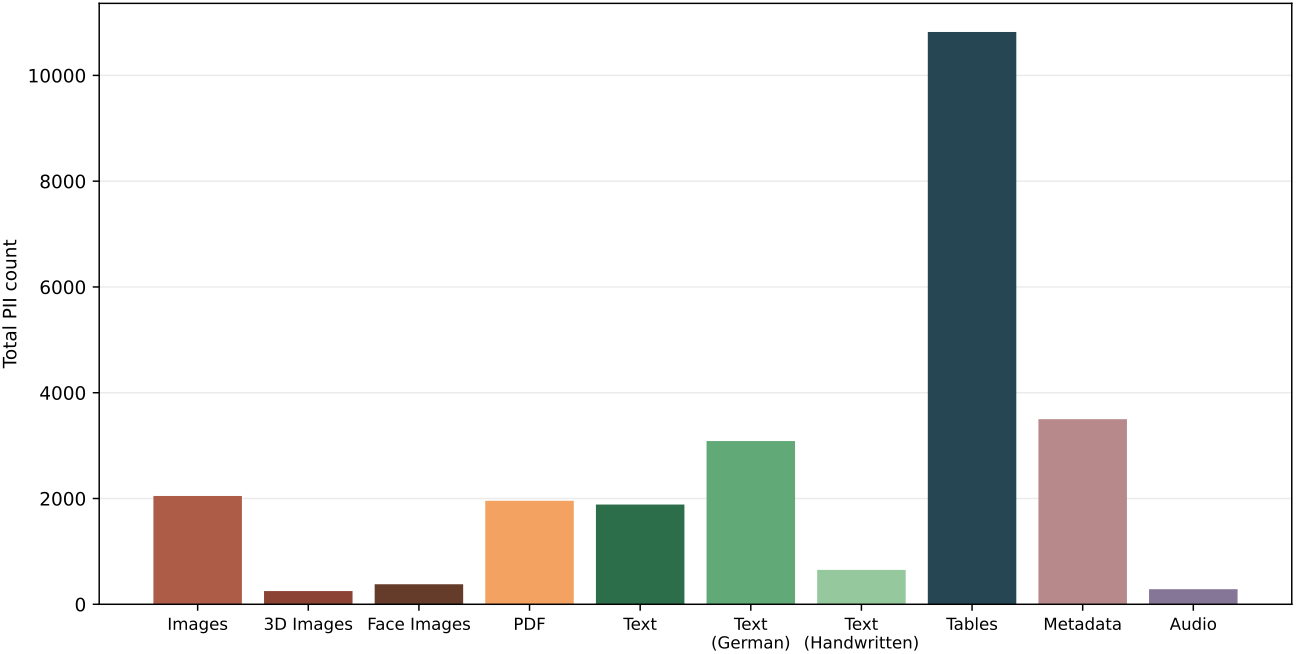
Total count of personally identifiable information (PII) instances across modalities in DeID-Extended.

**Figure S26:**
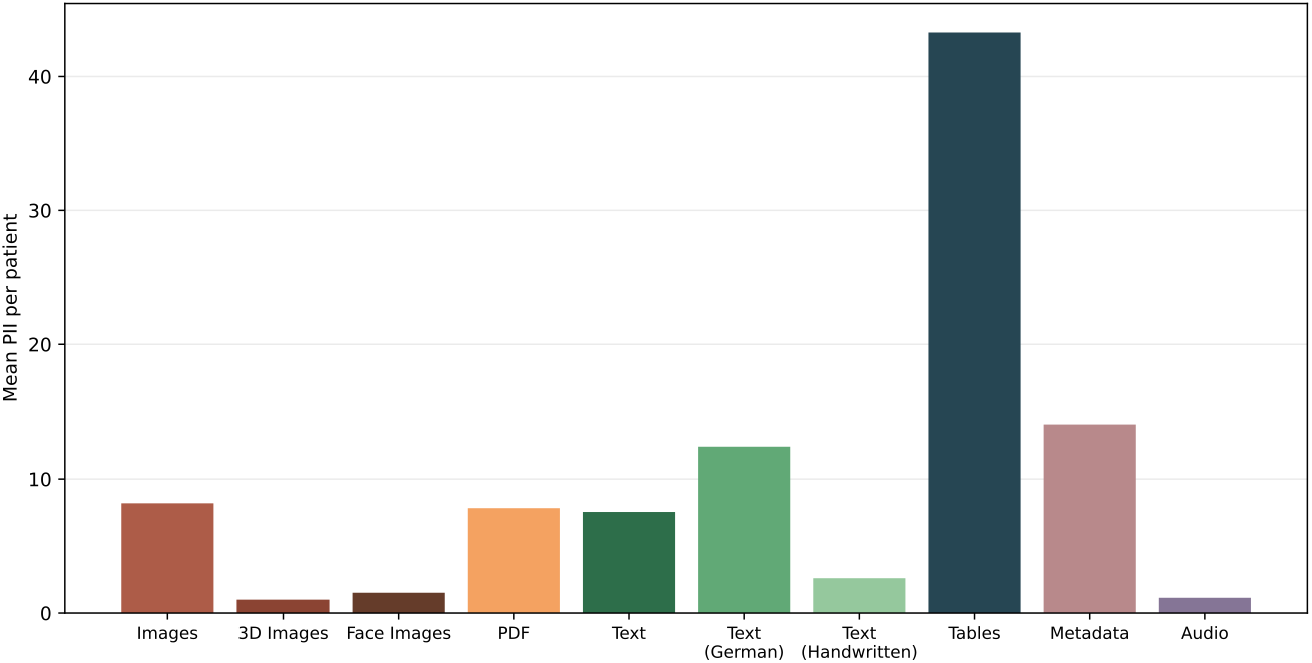
Mean number of personally identifiable information (PII) instances per patient across modalities in DeID-Extended.

**Figure S27:**
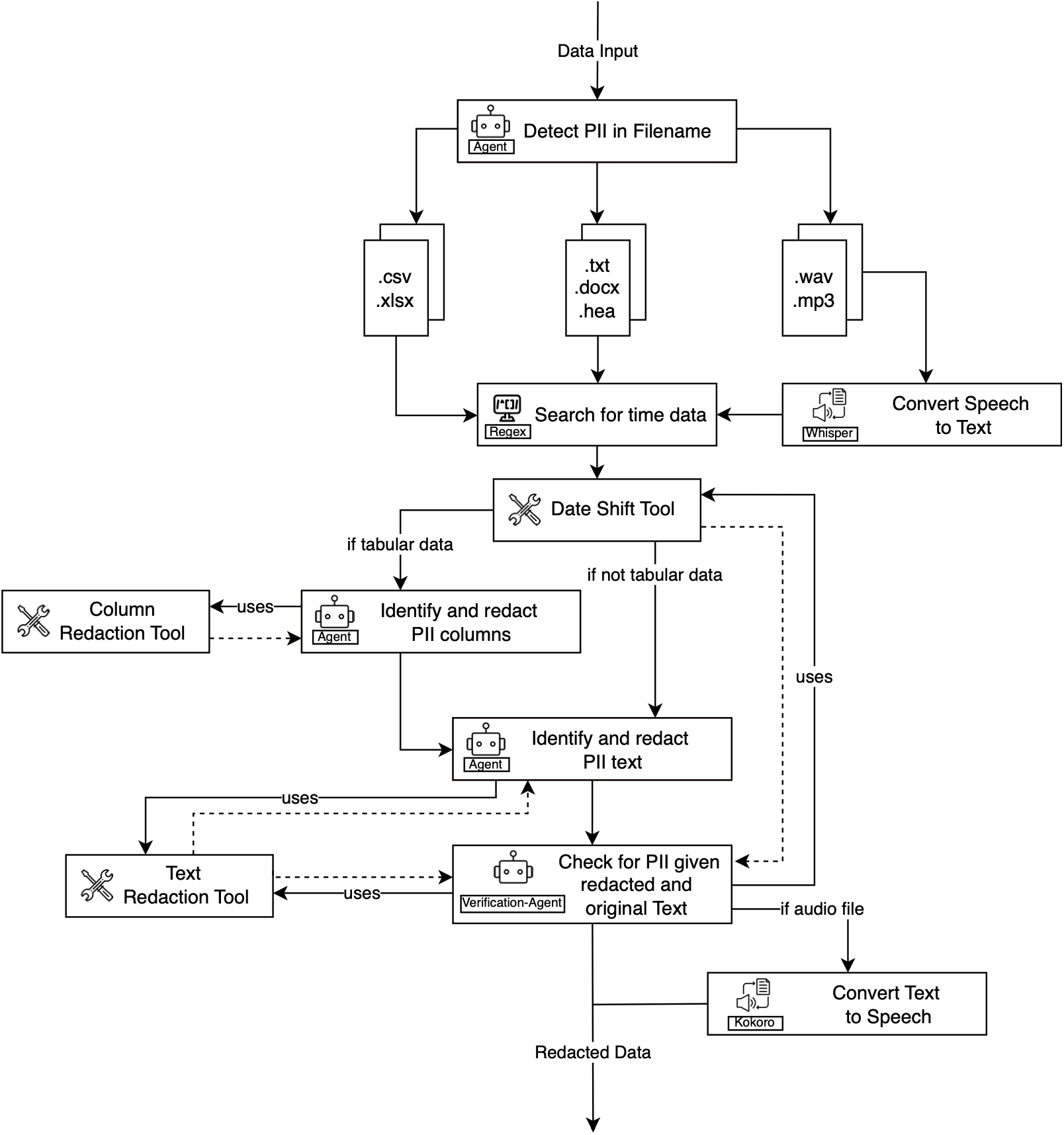
Multimodal Anonymizer Pipeline for textual data and audio. The pipeline begins by detecting and redacting PII in filenames using an MLLM; the files are then routed by type and processed with the corresponding processor: tabular data, text documents, and audio files. Audio files are first transcribed to text using whisper-large-v3*^5^*, after which all text-based content passes through a regex-based date detection step followed by a date shift (random number of days within ± 3 years) tool that applies consistent temporal offsets. For tabular data, a dedicated agent identifies and redacts PII-containing columns using a column redaction tool, while all text content is processed by a separate agent that identifies and replaces PII with standardized placeholders. A verification agent then reviews the redacted output (max. 3 times) against the original text to catch any missed identifiers, using the text redaction tool for corrections as well as the date shift tool if it detects an unshifted date. After the redaction for audio files, the text is resynthesized into speech using kokoro-onnx*^6–8^*and redacted spans are replaced with an acoustic masking signal.

**Figure S28:**
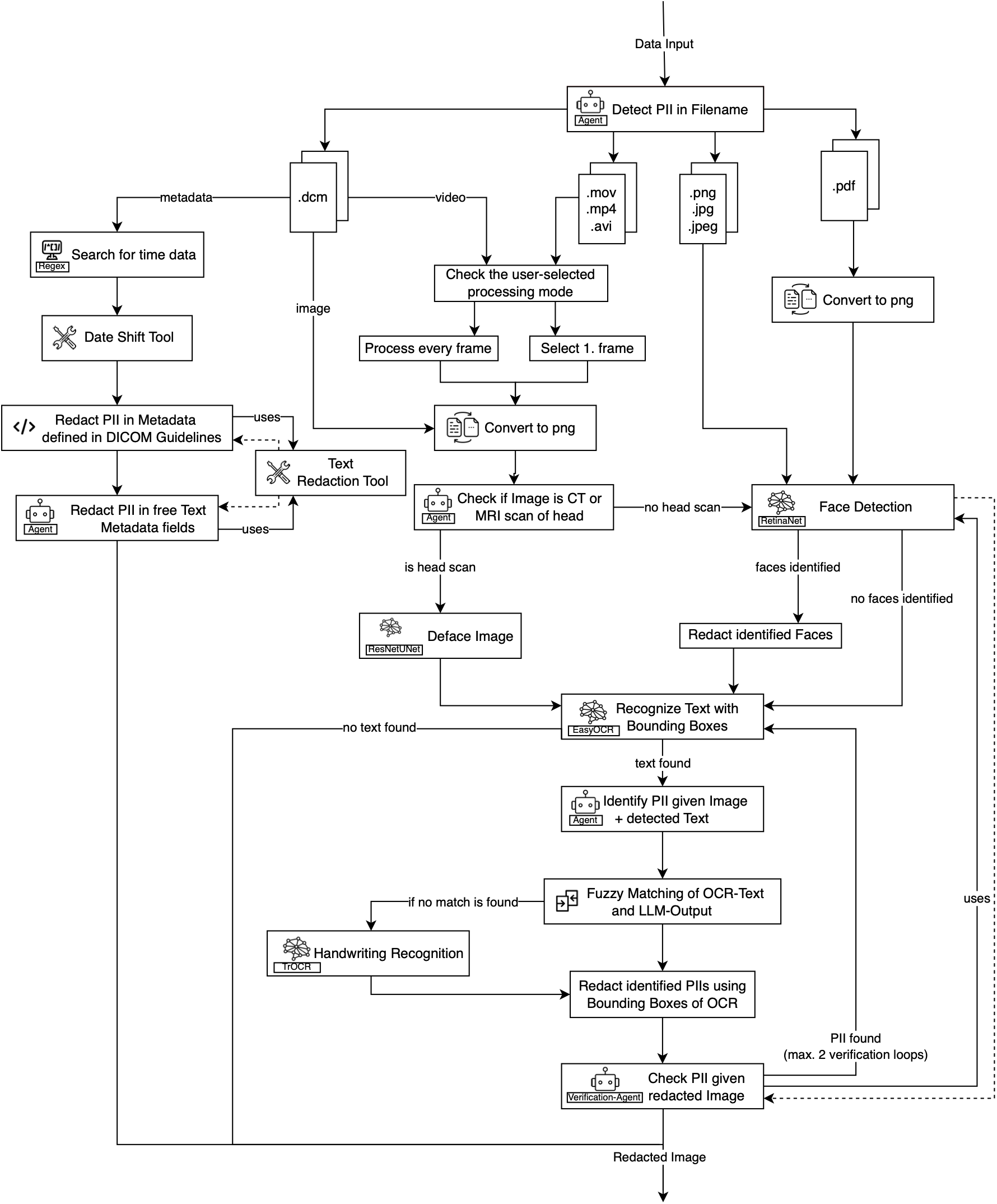
Multimodal Anonymizer Pipeline for images, PDFs, videos and DICOM files. After filename-level PII detection, the pipeline routes files by type. DICOM files are split into metadata and image components. Videos are processed either frame by frame or using only the first frame, depending on user preference; images are passed directly. For DICOM metadata, dates are identified via regex and shifted using the date shift tool, rule-based redaction removes identifiers defined in DICOM guidelines, and an MLLM agent redacts PII in free-text metadata fields. For all modalities, including DICOM, videos, images and PDFs, a temporary PNG representation is generated to be able to send the image to the MLLM. DICOM files and videos then pass through an MLLM agent that determines whether the image represents a volumetric head scan: if so, a ResNet-UNet model performs defacing (train dataset: size ∼1000 examples containing faces and ∼15000 Echo and CXR images as negative examples; test dataset: ∼300); otherwise, a RetinaNet model performs face detection (train dataset: size ∼1400 examples containing faces and ∼15000 Echo and CXR images as negative examples; test dataset: size ∼3500) and redacts any identified faces. All other modalities are likewise processed through RetinaNet for face detection. EasyOCR*^9^* then extracts burned-in text with bounding box coordinates from the original data, and an MLLM agent receives both the temporary PNG and the detected text to identify which elements constitute PII. Fuzzy matching aligns the MLLM-identified PII with the OCR bounding boxes to account for recognition errors; if matching fails, which can especially occur with handwritten content, a fine-tuned TrOCR model*^10,11^* (train dataset: size ∼93000; test dataset: size ∼23000) is invoked as a fallback to re-extract the text before reattempting alignment. All redactions are applied to the original file rather than to the temporary PNG representations. A verification agent then reviews the redacted output for residual identifiers, repeating the detection–redaction cycle for up to two iterations. For videos processed in “first-frame-only mode”, redactions identified on the initial frame are propagated to all subsequent frames.

**Table S5.**
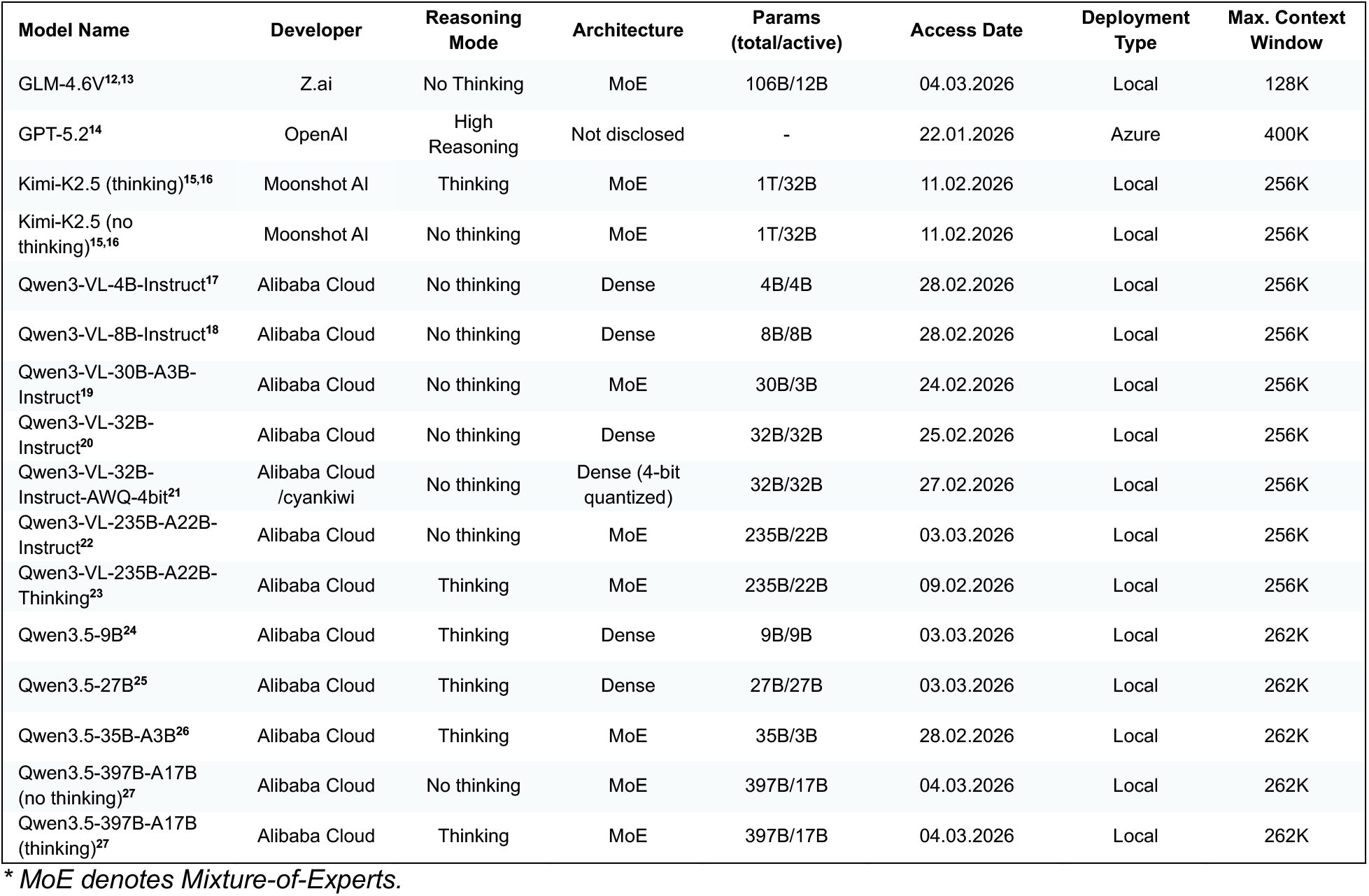
Model Configuration Details.*

**Figure S29:**
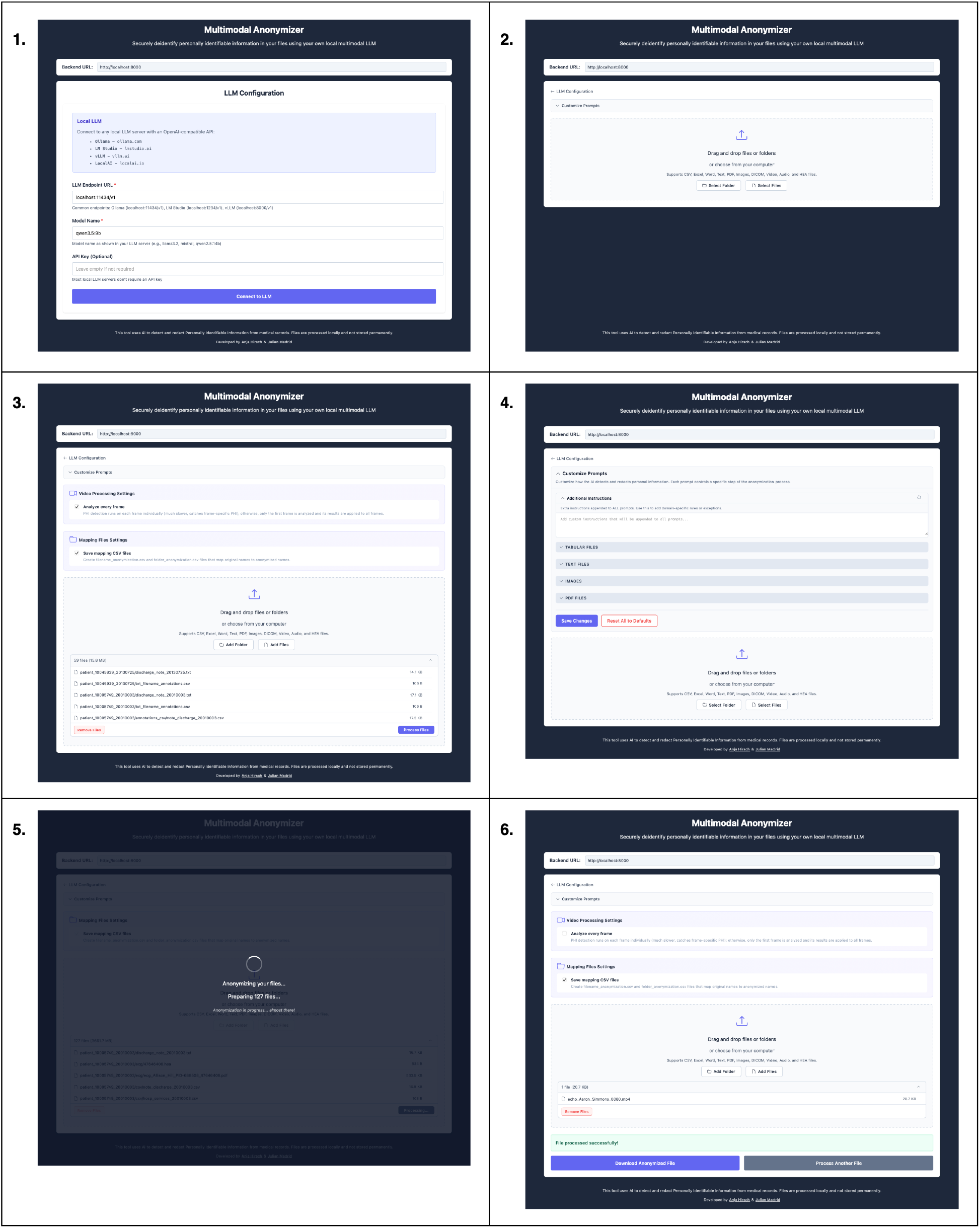
Multimodal Anonymizer Usage. The Multimodal Anonymizer is a browser-based application with two views and different functionalities. In addition, it can also be operated through the command-line interface (CLI). All instructions to set up the application are provided within the GitHub repository’s README file (https://github.com/charite-iaim/multimodal-anonymizer). After initializing the project, the following workflow within the User Interface can be used to perform the anonymization: **1. Model Configuration:** After starting the application, the user selects the LLM to be used for anonymization by providing a model endpoint URL and the model’s name. If the model requires an API key, it can be entered in the same form (though locally deployed models typically do not require one). Upon clicking "*Connect to LLM*", the application verifies whether a connection to the specified model can be established. If the connection fails, a descriptive error message is displayed; if successful, the user is directed to the next view, where files can be uploaded for processing. **2. File Upload:** Files and folders can be uploaded via drag and drop or by selecting them through the system’s file explorer. Uploaded items can be removed individually or cleared all at once. To modify the model configuration, the user can return to the previous view by clicking on "*LLM Configuration*". **3. Anonymization Configuration:** The user can optionally enable pseudonymization, which saves a CSV file mapping the anonymized filenames to their originals. If a video file has been uploaded, an additional option appears allowing the user to choose whether each frame should be processed individually or whether only the first frame should be anonymized and the redactions applied to all subsequent frames. **4. Prompt Customization (optional):** Prompts can be customized by clicking "*Customize Prompts*". The user can define an additional instruction that is applied globally to all prompts sent to the language model during the anonymization process. Beyond this, individual prompts for specific modalities and processing steps can also be adapted. For example, users can modify the prompt governing column-level redaction in tabular data or the prompt used during the verification phase. **5. File Processing:** Processing duration primarily depends on the inference speed of the underlying local LLM, making hardware configuration and model size key factors for overall throughput. **6. File Download:** Once all files have been processed, the anonymized results can be downloaded by clicking "*Download Anonymized Files*". When multiple files were processed, the data are provided as a compressed zip archive. If the pseudonymization option was selected, the CSV file mapping original to anonymized filenames is included in the download. To anonymize additional data, the user can click "*Process Another File*" to return to the upload view.

**Figure S30:**
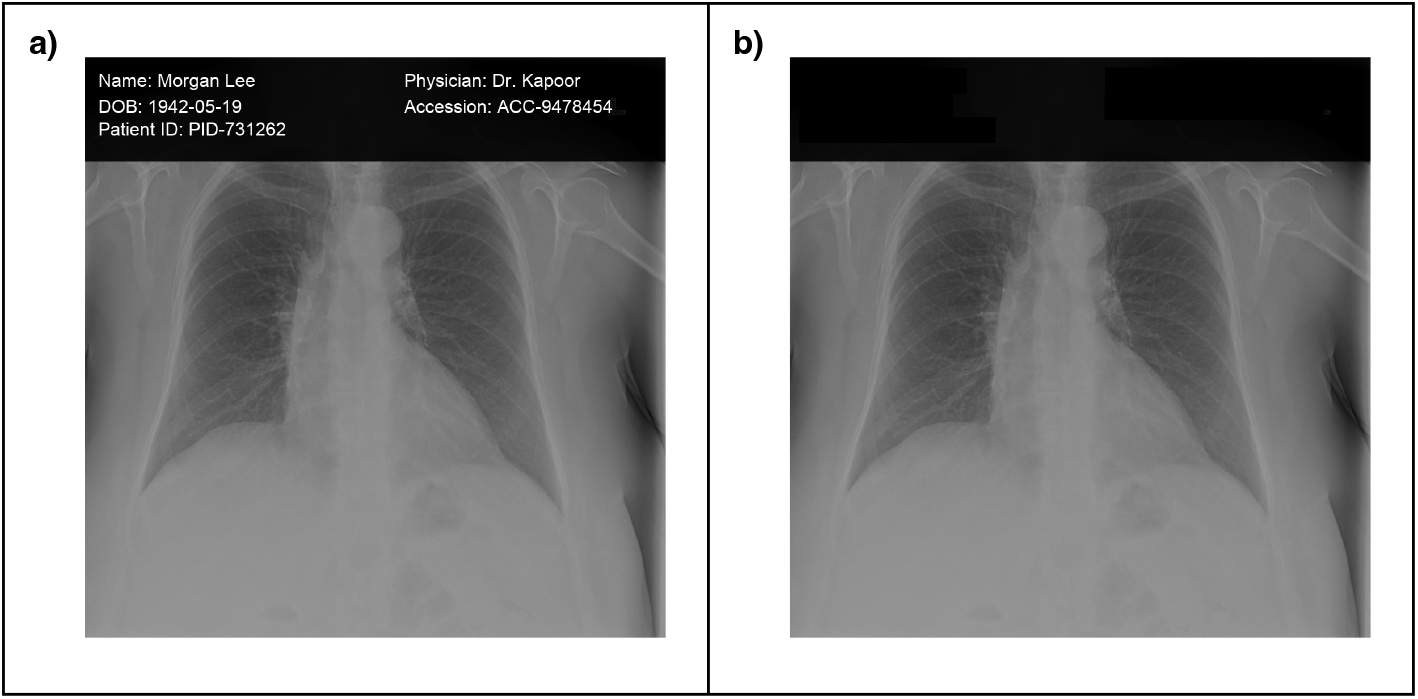
Results – Deidentification of a chest X-ray image. (A) Chest X-ray from the RSNA Pneumonia Detection Challenge dataset*^28^* with injected synthetic personally identifiable information. (B) Resulting image after anonymization with the Multimodal Anonymizer (Qwen3-VL-235B-A22B-Thinking).

**Figure S31:**
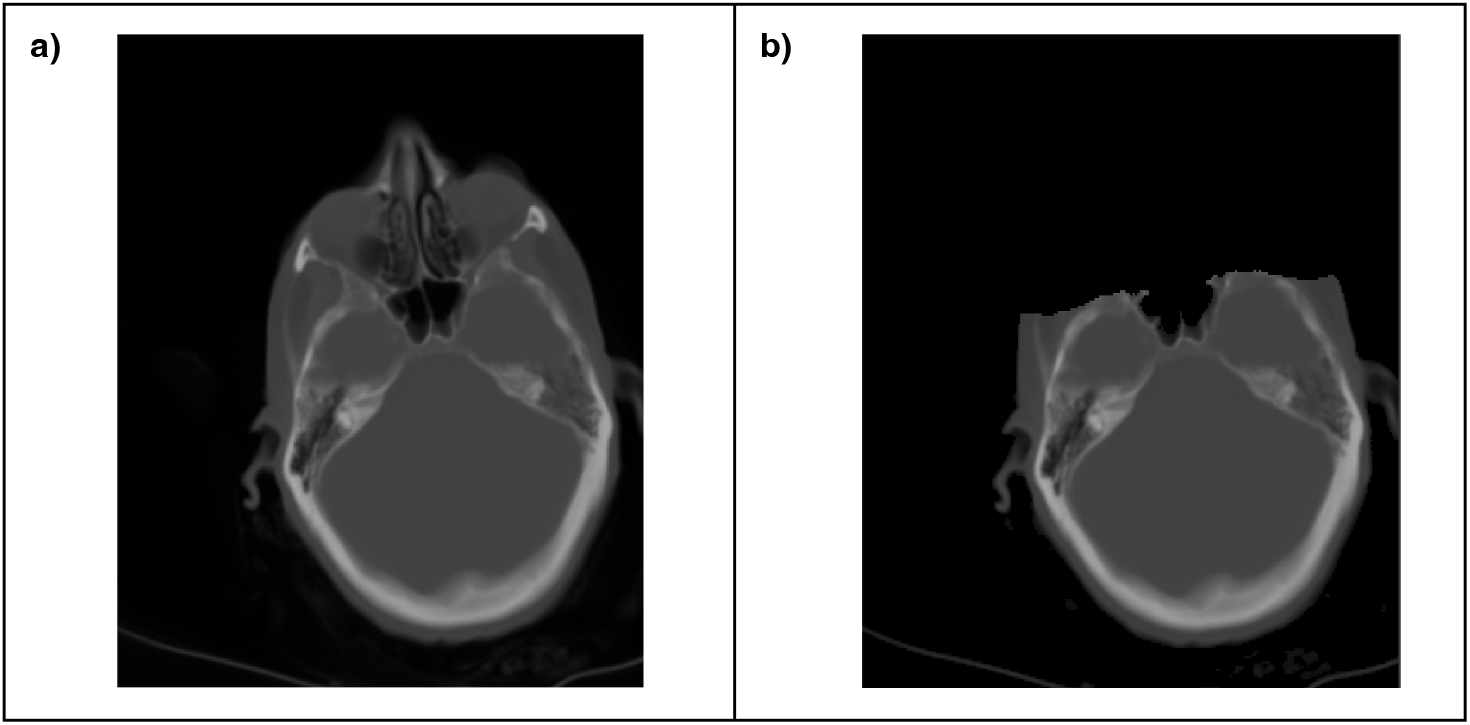
Results – Deidentification of a head CT image. (A) Axial head CT slice from a publicly available dataset*^29^*. (B) Resulting image after anonymization with the Multimodal Anonymizer (Qwen3-VL-235B-A22B-Thinking).

**Figure S32:**
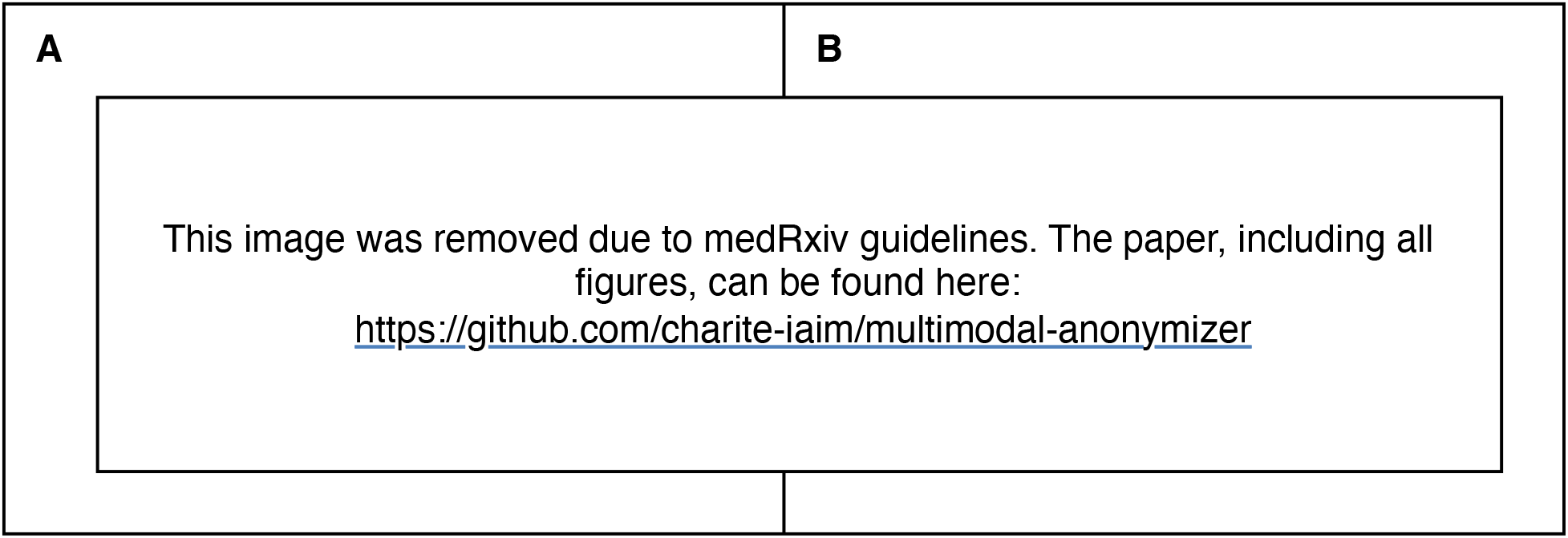
Results – Deidentification of a face image. (A) Image of a person with a face visible (image obtained at Charité – Universitätsmedizin Berlin; written informed consent for publication was obtained from the depicted individual). (B) Resulting image after anonymization with the Multimodal Anonymizer (Qwen3-VL-235B-A22B-Thinking).

**Figure S33:**
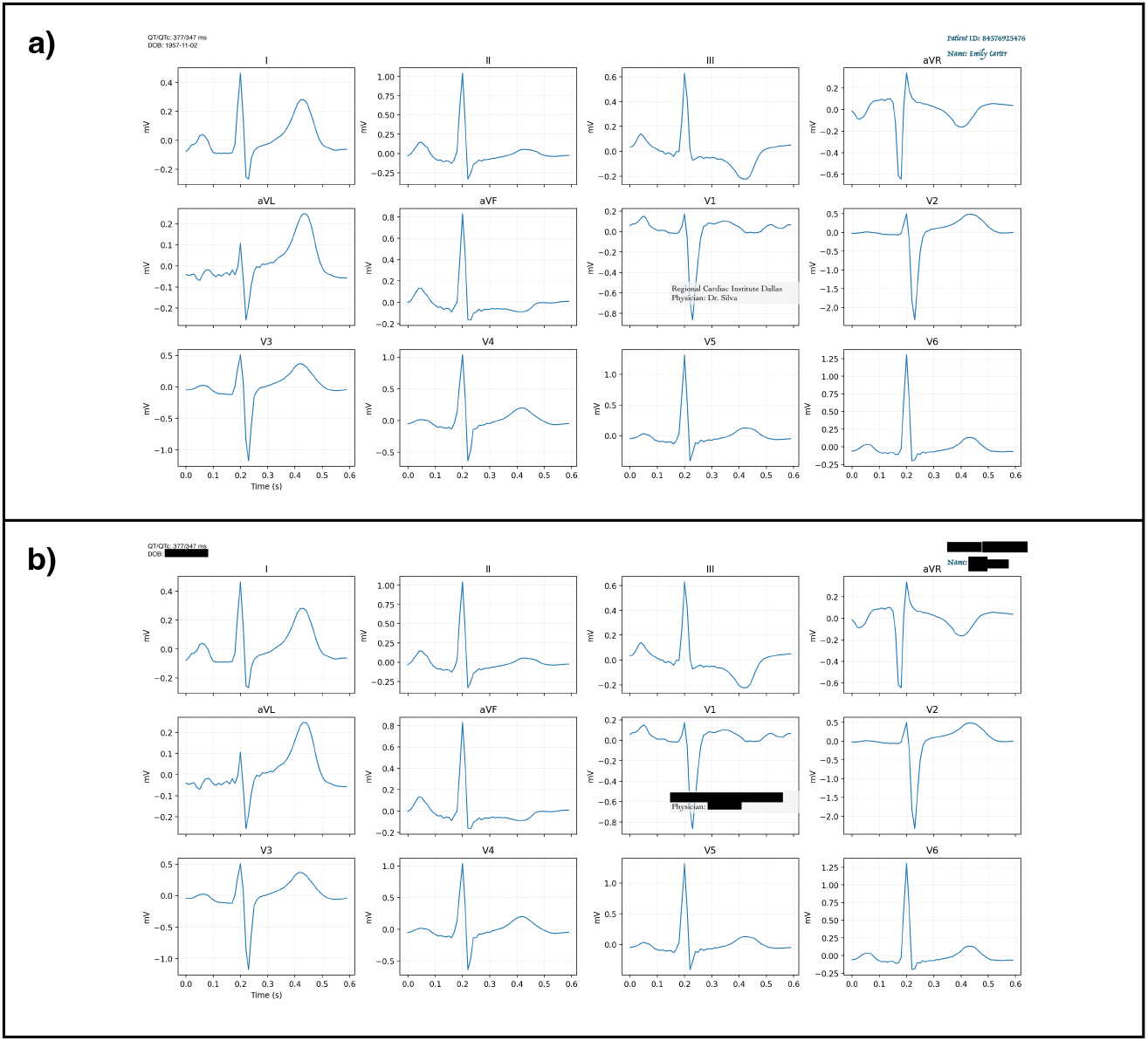
Results – Deidentification of an ECG. (A) ECG from the PTB-XL dataset*^30^* with injected synthetic personally identifiable information. (B) Resulting image after anonymization with the Multimodal Anonymizer (Qwen3-VL-235B-A22B-Thinking).

**Figure S34:**
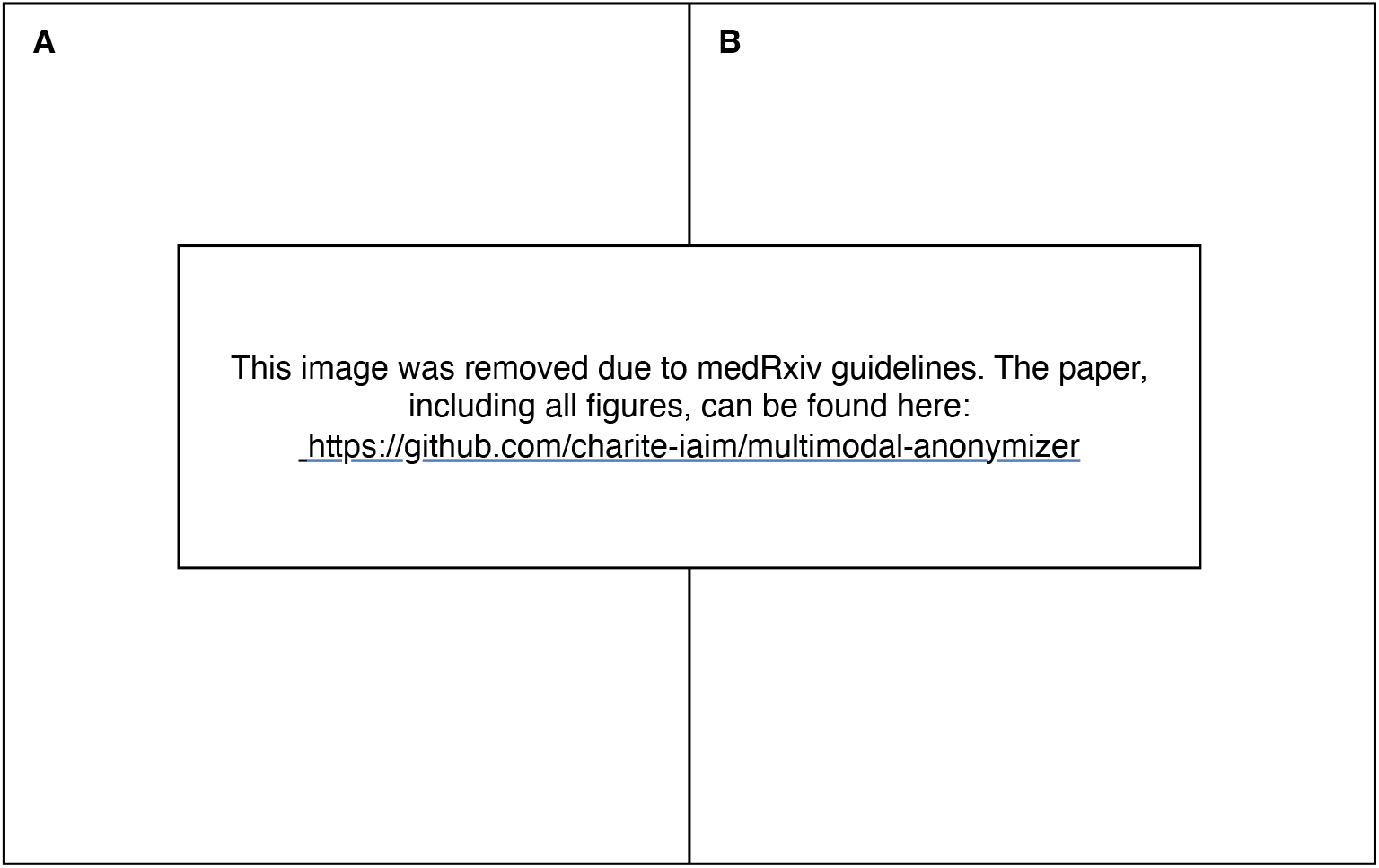
Results – Deidentification of a clinical letter. (A) German-language clinical letter from the GRASCCO dataset*^31^* containing synthetic personally identifiable information. (B) Resulting image after anonymization with the Multimodal Anonymizer (Qwen3-VL-235B-A22B-Thinking).

**Figure S35:**
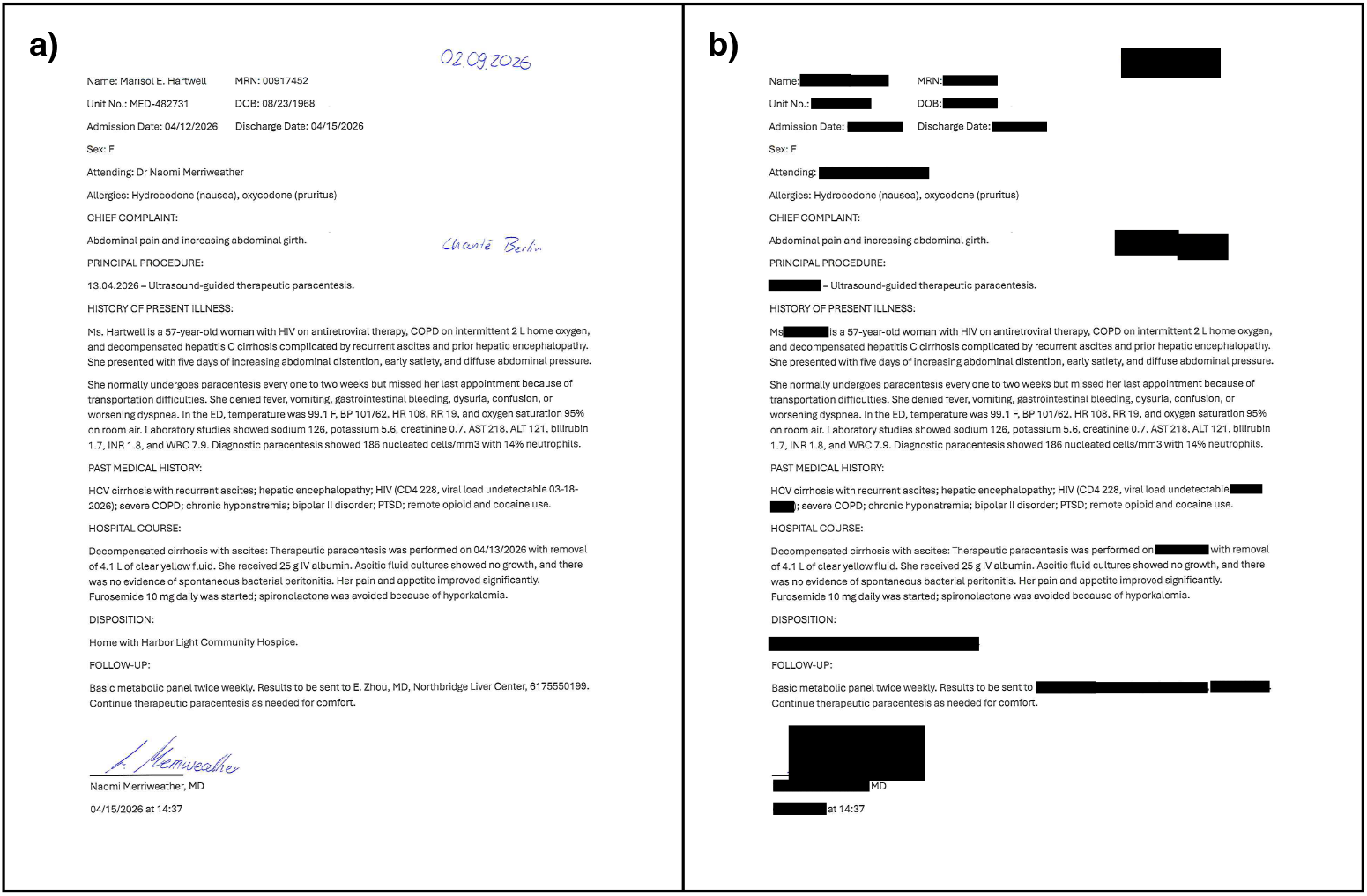
Results – Deidentification of a clinical discharge summary. (A) Fully synthetic clinical discharge summary with synthetic personally identifiable information, including handwritten annotations. (B) Resulting image after anonymization with the Multimodal Anonymizer (Qwen3-VL-235B-A22B-Thinking).

**Figure S36:**
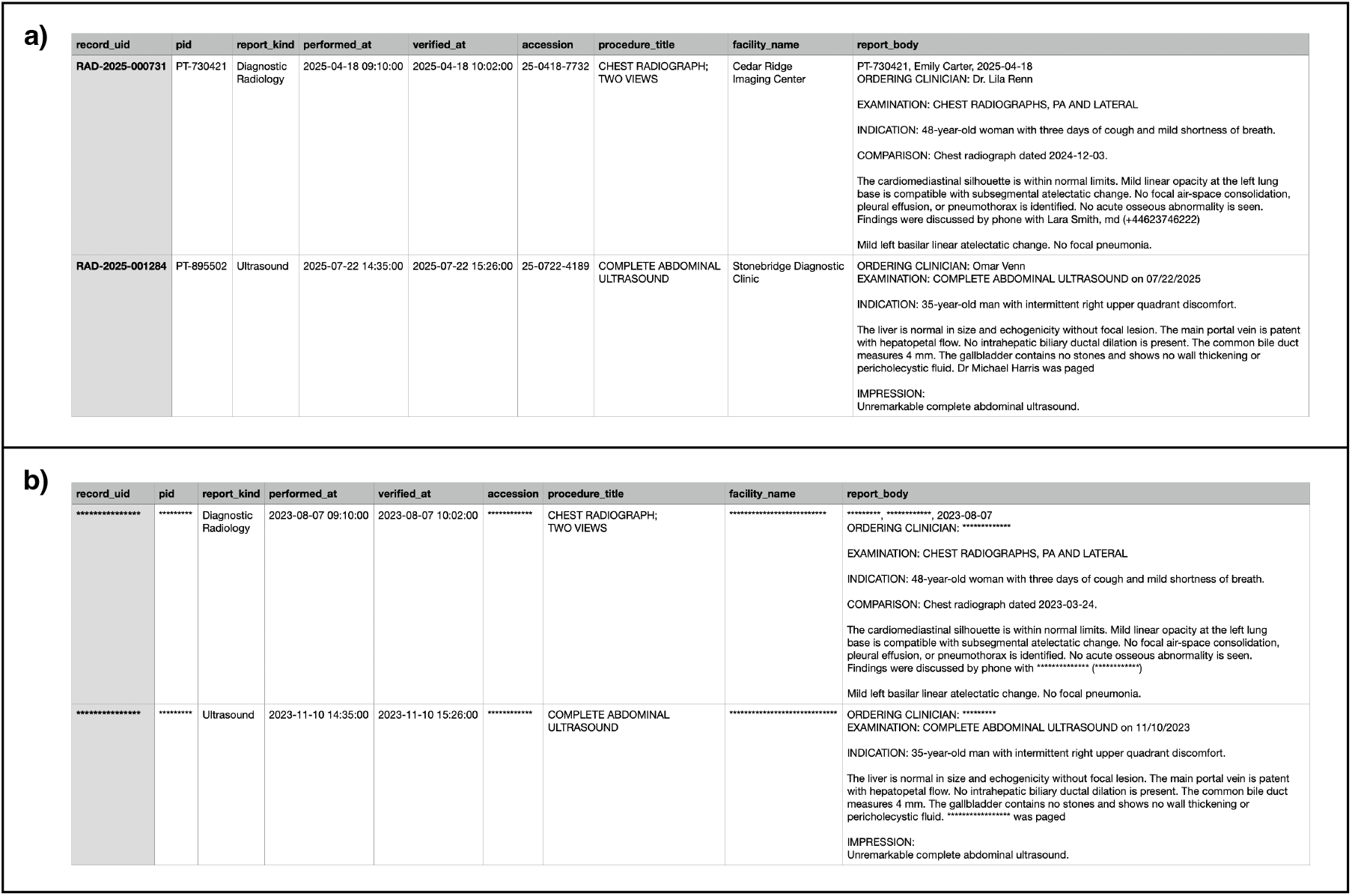
Results – Deidentification of a table. (A) Fully synthetic clinical table containing synthetic personally identifiable information across multiple fields. (B) Resulting table after anonymization with the Multimodal Anonymizer (Qwen3-VL-235B-A22B-Thinking).

**Figure S37:**
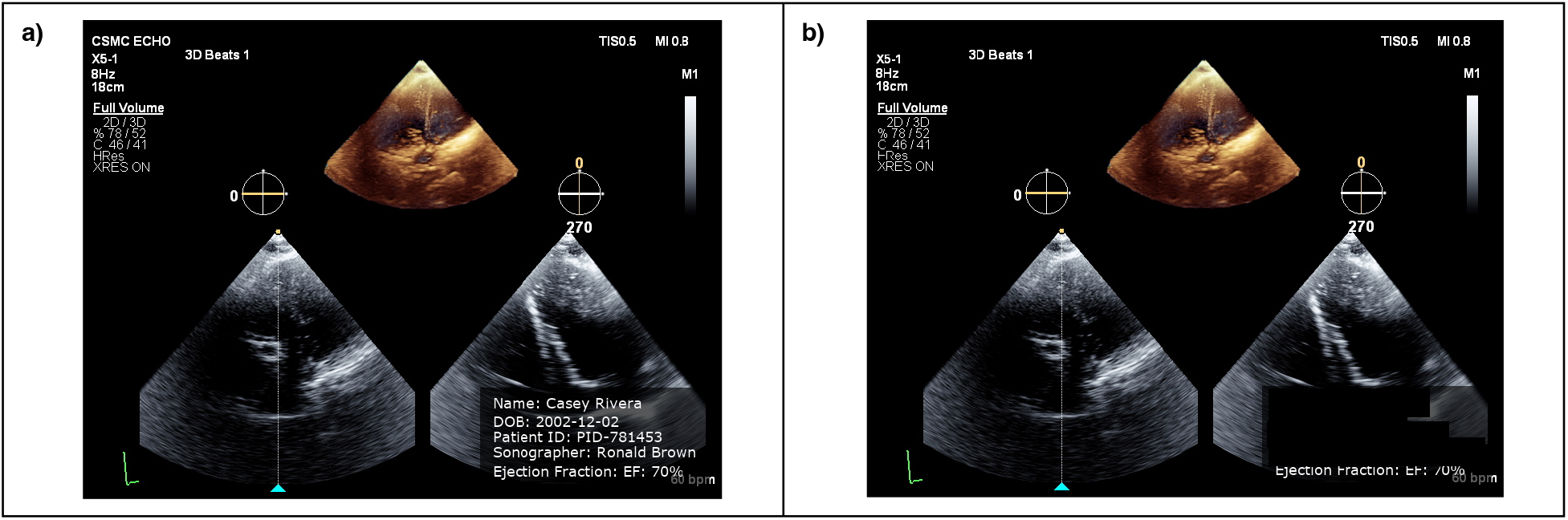
Results – Deidentification of an echocardiogram. (A) Echocardiographic image from the 3D echocardiography dataset accompanying EchoSlicer*^32^* with synthetic personally identifiable information injected into the DICOM file. (B) Resulting image after anonymization with the Multimodal Anonymizer (Qwen3-VL-235B-A22B-Thinking).

## Notes

### Competing Interest Statement

This study was supported by the Berlin Institute for the Foundations of Learning and Data (BIFOLD). AH, RG, AM, and JM received support from BIFOLD during the conduct of the study. TR and MIG were supported by the BIH Digital Clinician Scientist Program of the Berlin Institute of Health at Charite. The funders had no role in the design or conduct of the study; the collection, management, analysis, or interpretation of the data; the preparation, review, or approval of the manuscript; or the decision to submit the manuscript for publication.

### Author Declarations

Publicly available datasets were used in accordance with their respective access and licensing terms. The Charite Ethics Committee approved the retrospective use and anonymization of the institution-specific partogram dataset from Charite Universitaetsmedizin Berlin (EA2/267/25) without requiring individual informed consent. Reproducibility materials are provided as permitted by data-use agreements, institutional policy, and patient privacy requirements.

### Summary of Updates

The manuscript title has been revised to better reflect the scope and objectives of the study. The descriptions of the datasets have been expanded to provide greater clarity regarding the data sources, included modalities, cohort composition, and data preparation procedures. The Methods section has also been substantially revised to provide a more detailed description of the Multimodal Anonymizer workflow, including modality specific processing, anonymization procedures, and the evaluation methodology. Additional methodological details have been added to improve transparency and reproducibility, including clarification of the reference standards, evaluation procedures, and performance assessment. The manuscript has also been revised throughout to improve clarity, consistency, and the description of the overall study design.

